# Nanoparticle enriched mass spectrometry proteomics in British South Asians identifies novel variant-protein-disease mechanisms

**DOI:** 10.1101/2025.08.12.25333522

**Authors:** Maik Pietzner, Alice Williamson, Karen A. Hunt, Mine Koprulu, Leonhard Kohleick, Kamil Demircan, Genes & Health Research Team, Sarah Finer, Julia Carrasco Zanini, David A van Heel, Claudia Langenberg

## Abstract

Understanding genetic variation underlying differences in plasma protein levels can elucidate human disease mechanisms, but prior evidence was almost entirely derived from white Europeans using protein-preselected affinity reagents. Here, we integrate exome sequencing and common non-coding variation with untargeted nanoparticle enriched mass spectrometry (MS)-based plasma proteomics (Seer Proteograph XT: n=8067 protein groups) in >1,400 British-Bangladeshi and British-Pakistani individuals. We gain quantitatively and qualitatively different insights compared to two affinity-based assays in the same samples (SomaLogic 11k: n=9685 and Olink HT: n=5416 proteins), both in terms of proteins covered, and new proteogenomic insights into disease biology. Considering both additive and non-additive genetic effects, we identify >1,200 significant variant-protein associations (n=895 *cis*-protein quantitative trait loci (pQTL)), half of which are novel. Cross-platform comparison demonstrated that inconsistencies in pQTL discovery are mostly explained by technical variation, and that multiple platforms are required to capture the full spectrum of pQTLs of blood proteins. We integrate proteogenomic evidence with orthogonal human genetic, experimental, and single cell expression data to consolidate a potential role of 21 proteins in the pathology of 44 diseases: e.g., a novel role of high IGLV3-21 in the development of Grave’s disease elucidating B-cell mediated autoimmunity. Our results demonstrate the potential of MS-based blood proteomics in diverse ancestries for pQTL discovery, including functional characterization of missense variants, and the need to consolidate evidence from multiple biological domains to confidently assign proteins to disease pathology to guide drug target identification and drug repurposing.

## INTRODUCTION

Proteins circulating in blood, either actively secreted or leaked because of cell turnover/damage, have been shown to be under genetic control^1^. Such protein quantitative trait loci (pQTL) can inform drug target selection or drug repurposing in humans^2^. The most comprehensive studies so far used affinity-based methods, using either antibodies^3–6^ or single stranded DNA aptamers^7–13^, to measure up to ∼5000 preselected protein targets. However, between one-third^3^ and half^13^ of the assayed protein targets could not be linked to variation near or within the corresponding protein coding gene in studies of >30,000 participants although predictions indicated that those associations should have already been observed with 6-fold smaller sample sizes^3^. Among individual contributing factors specific to each protein target, missing biological detectability in blood or insufficient specificity or sensitivity of affinity-based reagents are the most likely reasons for this discrepancy^14,15^.

Plasma proteomic studies utilizing mass spectrometry (MS) previously measured 500 - 1200 proteins readily detectable in blood at moderate sample size, but quantified proteins and associated genetic variants not detected in much larger affinity-based efforts^16–19^. However, the large dynamic range of the plasma proteome that spans more than ten orders of magnitude^20^ provides a challenge for broader coverage of the plasma proteome, specifically low-abundant proteins with signalling capacity, like cytokines or hormones. While depletion and enrichment methods have been constantly developed during the past decades^20^, it is only now through the advent of nanoparticle-based enrichment and next generation MS-instruments that >5000 proteins circulating in blood can reproducibly be measured at scale^21,22^ providing unique opportunities for translational research.

Here, we present the most comprehensive MS-based plasma proteomic study so far, studying plasma levels of >5,700 protein groups (i.e., groupings of proteins that share identified peptides) robustly measured (>70% detection rate) in >1,400 volunteers of the Genes & Health (G&H) cohort of adult British Bangladeshi and British Pakistani ancestry participants^23^. These communities are at disproportionally high risk for cardiometabolic and other diseases^23^ and have been historically underrepresented in genetic studies despite its higher frequency of homozygous carriers of alternative alleles due to high rates of parental relatedness (autozygosity)^24^. We demonstrate important quantitative and qualitative differences in the coverage of the plasma proteome and subsequent genetically guided prioritization of disease-mediating proteins compared to affinity-based efforts. For example, we detect robust evidence for abundant missense variation, including variants >10-fold enriched in our population, affecting plasma protein levels not resolvable with affinity reagents. Our results suggest that pQTL discovery using MS in underrepresented ancestries provides an efficient strategy that is synergistic to ongoing biobank-scale efforts.

## RESULTS

We performed the most comprehensive MS-based plasma proteomic study to date, identifying a total of 8079 protein groups detected at least once across 1612 plasma samples of recalled volunteers of the Genes & Health (G&H) cohort of British Bangladeshi and Pakistani ancestry participants (N=1464, mean age 43.4 years (12.2 s.d.), 55.9% females) (**Supplementary Tab. 1**). This included 5768 protein groups seen in 70% or more of the samples, demonstrating the depth and coverage of the MS-based plasma proteome.

### MS-based plasma proteomics covers distinct fractions of the blood proteome

We identified more than 3430 proteins (1941 detected in >70%) in our nanoparticle coupled MS-based assay that were not covered by the two currently most comprehensive affinity-based platforms, SomaLogic 11k (n=4533 proteins shared) and Olink HT (n=2529 proteins shared). This highlights a substantial fraction of the circulating plasma proteome that is currently largely unexplored (**Fig. 1a** and **Supplementary Tab. 1-3**). Notably, proteins detected by MS, but also covered using other platforms, showed only small fractions of those predicted to be secreted into blood^25^ (Seer Proteograph XT: 6.6%, SomaScan 11k: 7.2%, Olink HT: 9.3%) providing evidence that most of the proteins readily detectable in blood originate from diverse sources and mechanisms (**Fig. 1b**).

We observed that the differential coverage across all three technologies translated into shared and distinct pathways and tissue/cell-type contributions enriched among proteins targeted by each platform (**Fig. 1c and Supplementary Fig. 1-2**). Proteins detected by MS showed the highest number of mutually exclusive pathways (**Supplementary Fig. 1 and Supplementary Tab. 4**) and were, for example, enriched for processes specific to blood cells, like neutrophil degranulation (odds ratio: 1.89; p-value<9.7x10^-93^). In contrast, proteins captured by affinity-based platforms were most significantly enriched for unspecific immune signalling pathways (e.g., SomaLogic 11k: odds ratio: 1.27; p-value<3.6x10^-50^), while extracellular matrix proteins were enriched across all platforms (**Supplementary Fig. 1**). We corroborated these quantitative differences by profiling tissue or cell-type specificity using bulk tissue and single-cell mRNA expression data (**Fig. 1c**). For example, proteins likely specific to blood forming organs (9.8%, n=832) and gastrointestinal tissues (11.5%, n=982) were most prevalent among those measured with MS, whereas targeted affinity-based platforms showed higher fractions of proteins likely specific to the brain or sex-specific tissues.

**Figure 1.**
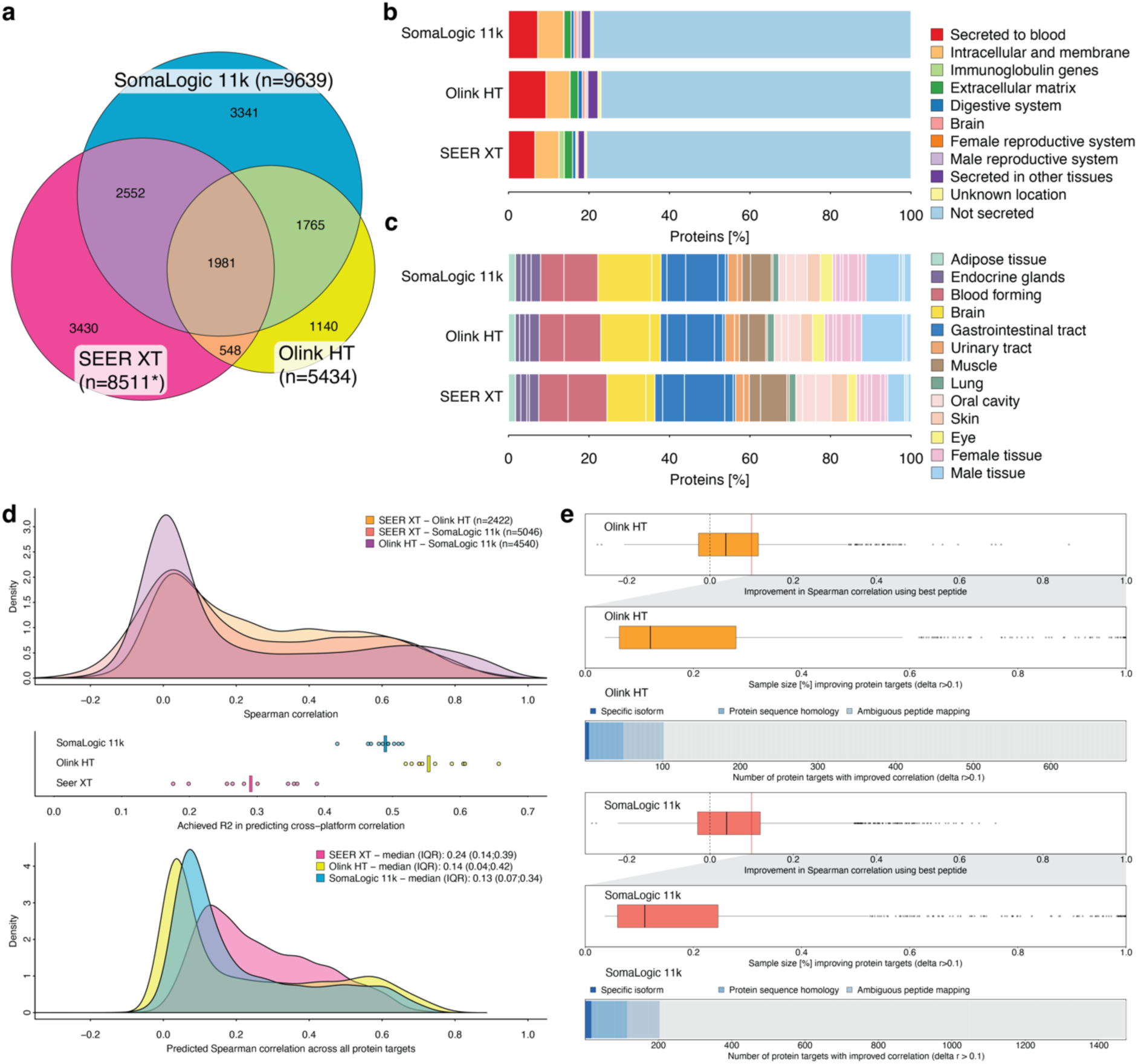
Quantitative and qualitative difference in mass spectrometry and affinity-based plasma proteomics. **a** Euler diagram showing the overlap of proteins captured by each platform (based on unique UniProt identifiers) *some protein groups mapped to multiple UniProt identifiers. **b** Assignment of secretome categories according to the Human Protein Atlas for each platform. **c** Fraction of protein targets with evidence for enhanced production in certain tissues (separated by lines) based on mRNA expression patterns reported by the Human Protein Atlas. **d** Distribution of Spearman correlation coefficients comparing measurements on the same samples for overlapping protein targets (top). The middle panel illustrates achieved R2 in predicting the best correlation value for each platform based on technical and other characteristics of the platform listed. Each dot is the result of one cross-validation step, and the bar indicates the median performance. The lower panel shows the subsequently predicted correlation coefficients even for targets unique to each of the platforms. **e** Comparing the best correlating MS-based peptide instead of protein groups with matching measurements from affinity-based efforts (Olink HT – top; SomaLogic 11k - bottom). Boxplots indicate the distribution of improvement (red line: delta r >0.1) in correlation and associated sample size for the respective peptide. Bar charts indicate potential explanations for improvements, if any.

### Technical variation explains poor cross-platform agreement of protein measurements and enables prediction of concordance

Plasma proteomic platforms have been reported to only partially agree^13–15,21,26^, which we replicate here across 6843 proteins overlapping across at least two platforms (**Fig. 1d** and **Supplementary Tab. 5-7**). We now demonstrate that the level of agreement across technologies can be largely explained (54% - 67%) by technical factors and host genetic information (**Supplementary** Fig. 3 and **Supplementary Tab. 8**). Building on that knowledge, we successfully trained three bespoke machine-learning models that utilized solely within-platform characteristics to predict the best correlation with any other assay (**Fig. 1d**). We observed that many protein targets are unlikely to achieve even weak cross-platform concordance (predicted r>0.1; **Fig. 1d**) when applying the models to the entire set of proteins covered by each technology irrespective of overlap. This included 13.5% (n=966; Seer XT), 45.5% (n=2453; Olink HT), and 39.1% (n=4194; SomaLogic 11k) of protein assays. These findings demonstrate the need to further improve assay performance and more specific selection of protein targets reliably detectable in blood in the general population.

Notably, considering peptide-instead of protein group-level MS measurements led to higher (delta r>0.1) correlation coefficients of plasma levels for 721 and 1401 proteins captured by Olink or SomaLogic, respectively (**Fig. 1e** and **Supplementary Fig. 4 and Tab. 5-7**). However, most of these improvements (83.1% and 88.8%) were only seen with peptides detected in less than half of the population (**Fig. 1e**) making it hard to distinguish from random improvements. We further obtained little evidence that protein isoform-specific targeting (n=5 antibody reagents; n=17 aptamer reagents), unspecific targeting of affinity reagents (n=44 antibody targets: n=97 aptamer targets), or ambiguities in bioinformatic peptide assignments to protein groups (n=71 antibody targets; n=125 aptamer reagents) explained poor correlations across protein platforms.

### Rare to common protein quantitative trait loci

We identified 1217 independent, significant (p<8.7x10^-12^) associations between 1016 genetic variants and plasma levels of 978 protein groups (out of 5768 detected in >70% of volunteers) by integrating genome-(minor allele frequency (MAF)>1%) and exome-wide (minor allele count >5) association results using statistical fine-mapping (**Fig. 2a** and **Supplementary Tab. 9**; see **Methods**). This included 94 variant – protein group pairs associated with >1 s.d. change per affect allele (MAF range: 0.2% – 41.0%). For example, the common variant chr18:12308794:G>C (MAF=12.7%) associated with 1.24 higher s.d. values for plasma levels of tubulin beta-6 chain, a protein involved in microtubule cytoskeleton organisation. More than 90% (n=1026) of the associations had support from three or more independent peptides, corroborating our strategy to minimize artificial findings by omitting genetically induced variant peptides from protein group quantification^17,19^ (see **Methods**). Notably, protein groups associated with genetic variants were almost 4-fold (odds ratio: 3.89; p-value<6.8x10^-38^) and 3-fold (odds ratio: 2.83; p-value<7.9x10^-8^) enriched for those predicted to be secreted to blood or the extracellular matrix, respectively, supporting the notion that proteins belonging to these compartments are most accessible for blood-based pQTL discovery.

Half of all identified associations (n=617) have not previously been reported (**Fig. 2a and Supplementary Tab. 9**), explained by the unique coverage of proteins by MS (n=460), but also including 13 loci never linked to plasma protein levels so far, demonstrating the potential of deep MS-based platforms to considerably expand the detection of pQTLs. This was even more apparent considering the 143 associations (n=42 close to the protein coding gene – ‘cis’) we identified at previously reported loci and for which we established novel protein associations despite the respective protein target being captured in studies up to 30 times larger than ours (**Fig. 2a**). A total of 17 unreported associations were most likely explained by strong differences in allele frequencies across ancestries, being common in our study of volunteers from South Asian ancestry but rare (MAF ≤ 0.5%) in the predominantly White-European ancestry participants of previous studies. For example, we identified a common synonymous variant (chr17:2042053:C>A; MAF = 10.0%) to be strongly associated with plasma levels of esterase OVCA2 (beta: -0.51; p-value< 2.1x10^-16^), which is almost absent in Non-Finnish European populations (gnomAD MAF = 0.01%). Irrespective of novelty, a total of 43 identified variants had effect allele frequencies >10-fold higher compared to White-European ancestries. For example, the missense variant chr14:105737776:C>T (p.G396R) in *IGHG1* associated with lower plasma levels of bromodomain adjacent to zinc finger domain 1B (BAZ1B; beta=-0.46; p-value<3.1x10^-20^) in trans, is frequent in South Asians (MAF=16.3%) but rare in White-Europeans (gnomAD MAF = 0.06%). The variant is also known as hIgG1-G396R and has been associated with susceptibility to systemic lupus erythematosus in individuals of East Asian ancestry^27^ conferring higher antibody production by B-cells, including autoantigens. BAZ1B is involved in chromatin remodelling and hence ubiquitously expressed, but no role in systemic lupus erythematosus or B-cell biology has been described so far.

We next sought to distinguish specific from pleiotropic regulatory variants (**Fig. 2b**) by clumping variant-level associations across the proteome into 898 pQTLs. We found that most pQTLs likely acted on the respective protein coding gene in proximity (±500kb; n=794 cis-pQTL), including 29 cis-pQTLs being associated with two and up to three distinct protein groups. For example, a pQTL proxied by the two common variants, chr19:38326336:T>C and chr19:38304610:G>A (r^2^=0.6), was significantly associated with lower levels of protein phosphatase 1 regulatory subunit 14A (PP14A; beta = -0.48; p-value<1.8x10^-24^) and immortalization up-regulated protein (IMUP; beta = -0.46; p-value<9.7x10^-26^). While the association with PP14A has previously been reported^3,4,10,11,13^, the association with IMUP is unique to MS-based platforms due to missing coverage by previous affinity-based platforms. This demonstrates the need for additional coverage of the plasma proteome to guide downstream applications that use cis-pQTLs to infer causal relationships between proteins and diseases as in Mendelian randomization.

The remaining pQTLs (n = 104) associated with distally encoded (‘trans’) proteins (**Fig. 2b and Supplementary Tab. 9**), including 43 pQTLs with cis- and trans-associations, possibly suggesting a role of the cis protein in elucidating trans effects. For example, a cis-pQTL proxied by the upstream gene variant chr5:177413083:G>GC for factor XII (beta: 0.96; p-value<4.3x10^-132^) was associated with a total of 20 protein groups more than 23-fold enriched for members of the complement and coagulation cascade (fold-change: 23.8; p-value<2.2x10^-6^). A finding in line with the central role of factor XII activation in initiating the intrinsic pathway of the coagulation cascade responsible for blood clot formation following endothelial injury. In contrast, the 60 protein groups associated with the most pleiotropic trans-pQTL **(Supplementary Tab. 9**), proxied by a 3’-UTR variant for *NINJ1* (chr9:93122219:A>G), were not enriched for any pathway. The unspecific nature of the associations might be attributed to *NINJ1*’s role in programmed cell death and subsequent propagation of blood cell death during sample handling. Briefly, we^14^ have provided evidence that similar unspecific trans-pQTLs can be linked to blood cell characteristics, including susceptibility to external stimuli. NINJ1 is a cell surface protein that mediates plasma membrane rupture upon sensing of damage-associated molecular patterns (DAMPs). Bone marrow derived macrophages of *Ninj1^-/-^* mice showed a differential ‘secretome’ upon cell death^28,29^, which may explain genetically induced protein changes associated with blood cell contamination even among samples that have been treated with the same tight protocol (≤1 hour until freezing at -80°C in the present study).

### Protein altering variants and overlap with tissue gene expression loci explain pQTLs

We observed a high fraction of protein altering variants (PAVs; 22.4% and 39.9%, respectively; **Fig. 2c**) for both cis-(n=325; 38.8%) and trans-pQTL (n=34; 44.6%) when functionally annotating pQTLs or their proxies (r^2^>0.6). This observation is particularly noteworthy, since we excluded all variant peptides from protein quantification for which we had evidence for at least one carrier in our study population based on exome sequencing (see **Methods**). Missense variation can alter protein levels in multiple ways, including but not limited to, protein stability, secretion, mis-localisation, or degradation eventually resulting in lower or higher plasma levels^30,31^. About 30% (n=41) of the 144 cis-pQTLs mapping to PAVs recorded in the ProtVar database^32^ are predicted to likely destabilise (ΔΔG>2kcal/mol) the associated protein. For example, chr20:24971511:G>A (p.L163F) maps to a highly conserved residue of adipocyte plasma membrane-associated protein (APMAP), was associated with substantially lower plasma levels (beta=-0.99; p-value<1.6x10^-24^), and had a predicted ΔΔG=10.3 strongly suggestive of destabilising the protein. Reliably quantifying the effect of PAVs on protein abundance in blood is therefore of great importance but can be challenging using affinity-based techniques, since it can be hard to distinguish true changes in protein levels from changes in the binding affinity of reagents to the epitope^1,13–15^.

We further identified evidence for 651 pQTLs that those may act via altered gene expression in one or more tissues^33^, including 545 cis-pQTLs aligning with cis-eQTLs, as they were putative causal variants in gene expression credible sets (posterior inclusion probability (PIP)>10%; **Supplementary Tab. 9**). Combined with the effects of PAVs this pointed to the potential underlying mechanisms for 85.8% of all identified pQTLs (n=771).

**Figure 2.**
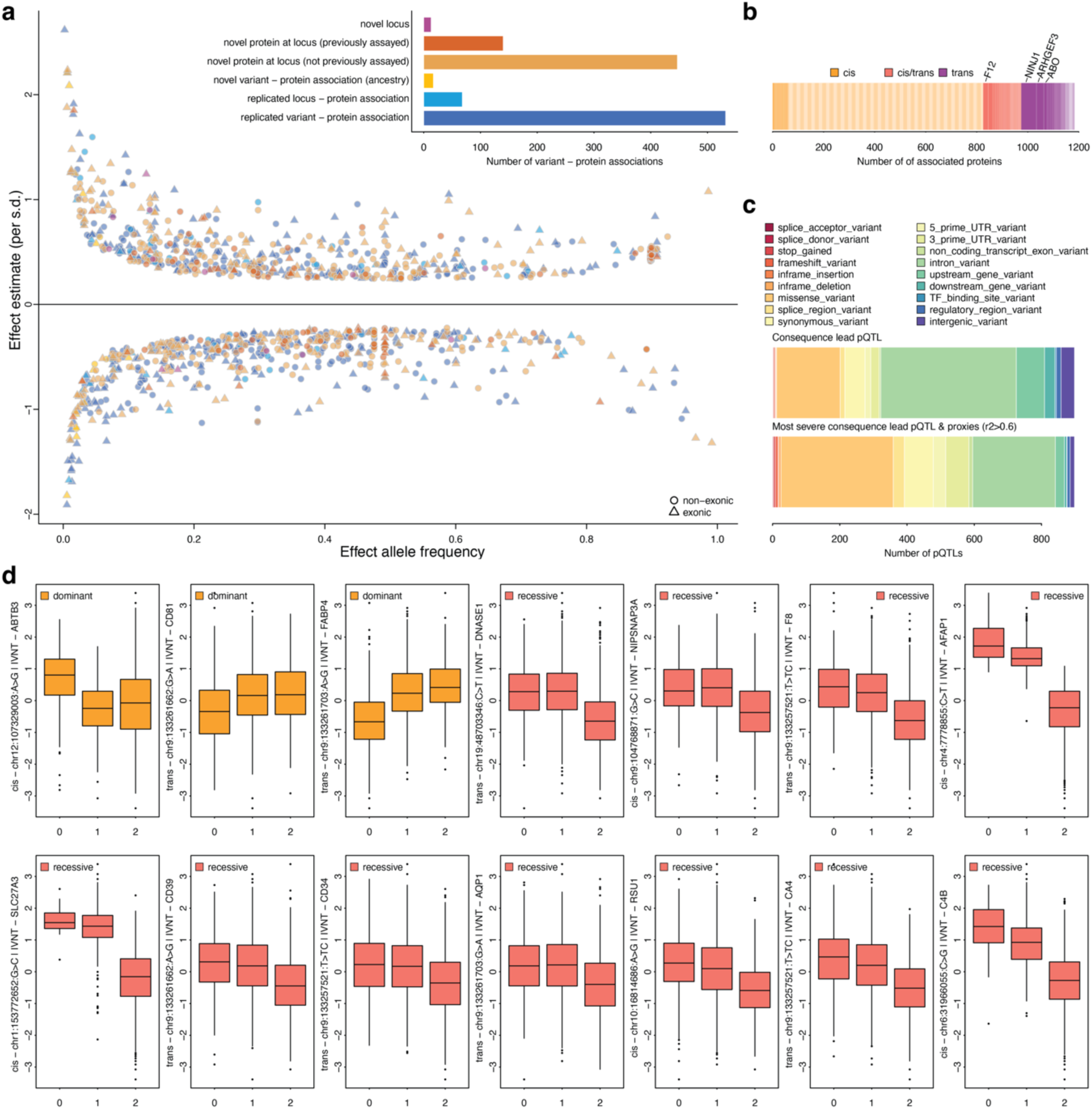
Rare and common pQTLs. **a** Scatterplot opposing effect allele frequency and effect estimate for associated genetic variants. Symbols indicate whether the variant is based on genotype-imputation(non-exonic) or whole exome sequencing. The inset quantifies the degree of novelty and is used as a colour gradient in the scatterplot. **b** Barchart displaying the number of pQTLs inferred from LD-based clumping across protein targets. pQTLs with strong evidence for pleiotropy have been annotated. **c** Barcharts showing the functional annotation of identified genetic variants or proxies thereof (lower panel). **d** Selected genetic variant – protein associations with evidence for non-additive genetic effects. Colour codes indicate the likely mode of inheritance based on the reference allele.

### Non-additive genetic effects for cis- and trans-pQTL

The enrichment of homozygous carriers of alternative alleles in the Genes and Health cohort (due to autozygosity) enabled us to systematically test for non-additive genetic effects across pQTLs. We identified robust evidence for autosomal dominant (n=3) or recessive (n=11) effects with respect to the reference allele, that were also supported by two or more peptides (**Fig. 2d and Supplementary Tab. 10**). We identified six non-additive effects close to the protein coding gene that have not been described to date. This included three examples for which homozygous carriers of the alternative allele had >1.5 s.d. units lower plasma protein levels compared to carriers of at least one reference allele (**Fig. 2d**). For example, homozygous carriers of the common G-allele for chr1:153772652:G>C had 1.64 s.d. units lower plasma levels of long-chain fatty acid transport protein 3 (FATP-3 or SLC27A3) compared to carriers of the minor C-allele. SLC27A3 activates long-chain fatty acids^34^ and a burden of rare loss of function or missense variants has previously been associated with lower asthma risk^35^. Non-additive effects in trans were further exclusively explained by recessive effects on proteins involved glycosylation, namely alpha 1-3-galactosyltransferase (ABO, e.g., via chr9:133257521:C>CT, n=7) and fucosyltransferase 2 (FUT2, via chr19:48703346:C>T, n=1), which are known to encode blood group status but possess a wide spectrum of protein targets.

### Cross-platform concordance of pQTLs

We^14^ and others^13,15,36^ have previously reported pQTLs specific to the platform used for the measurement of the protein target. We were able to test for replication of our MS-based discovery effort using two affinity-based assays in the same volunteers (n=510 and n=1009, genetic variant – protein group associations using Olink HT and SomaLogic 11k, respectively). We observed that only about half of the MS-based associations (53.5% for Olink HT and 43.5% for SomaLogic 11k) showed at least moderate evidence of replication (concordant effect sizes and p-value<10^-5^) with low correlation of effect estimates (**Fig. 3a and Supplementary Tab. 11**). Lack of replication was almost exclusively explained by poor correlation of the corresponding protein measurements (**Fig. 3b** and **Supplementary Fig. 5-6**). Accordingly, most non-replicating associations (Olink HT: 79.8%; SomaLogic 11k: 83.3%) included protein targets for which we could not identify any genome-wide significant (p<5x10^-8^) cis- or trans-pQTLs using affinity-based measurements. Among those without replication but with likely distinct significant cis-pQTLs, about a third associated with PAVs (18/42 based on Olink HT and 27/85 based on SomaLogic). This suggested that differential findings might, in part, be a result of affinity-based limitations in recognizing altering binding epitopes^14^.

Replication of 2757 affinity-based pQTLs (440 cis- and 221 trans-pQTL for Olink HT; 684 cis- and 1412 trans-pQTLs for SomaLogic 11k) *(Williamson et al. in preparation)* using MS results, revealed lower replication rates (44.0% Olink HT; 22.9% SomaLogic 11k) despite higher correlations of effect estimates for targets overlapping with the Olink HT platform (**Fig. 3c** and **Supplementary Tab. 12**). We note that almost half (45.1%) of non-replicating trans-pQTLs of the SomaLogic 11k platform were accounted for by 5 highly pleiotropic trans-pQTLs we previously related to specifics of the measurement technique, such as genetically conferred higher affinity of complement factor H for DNA possibly binding non-specifically to many aptamers and hence biasing measurements^14^.

Non-replicating pQTL associations were best explained by poorly correlating protein measurements across platforms even when considering dozens of variant and protein characteristics in machine learning-based feature selection (**Fig. 3d and Supplementary Fig. 7-8**). This suggests that technical factors account for most non-replicating pQTLs irrespective of the platform used. This also questions the feasibility of recently proposed MS-based scores to flag artificial affinity-based pQTL efforts^37^. For example, among the 81 pQTLs from the two so far largest pQTL efforts proposed to be invalid and captured in our data, we observed only 25 protein targets to correlate sufficiently well (r>0.5) between MS- and affinity-based measurements to allow for such inference (i.e., that both platforms measure the same protein). In general, the extent to which PAVs or alike possibly introduce artificial pQTLs when relying on affinity reagents seemed to be moderate. For example, we identified robust evidence for only 46 out of 1133 instances that correlation coefficients were statistically different according to genotypes of associated cis-pQTLs indicating genetically induced poor assay performance (false discovery rate < 5% and significantly different correlation coefficients p<4.4x10^-5^; **Fig. 3e** and **Supplementary Tab. 13**). For less than half (n=20) of the examples, we were able to identify PAVs to possibly explain such effects, i.e., that the altered protein sequence may infer with the recognition by the affinity reagent. Another explanation might be that those cis-pQTLs tag splicing QTLs producing different protein isoforms.

**Figure 3.**
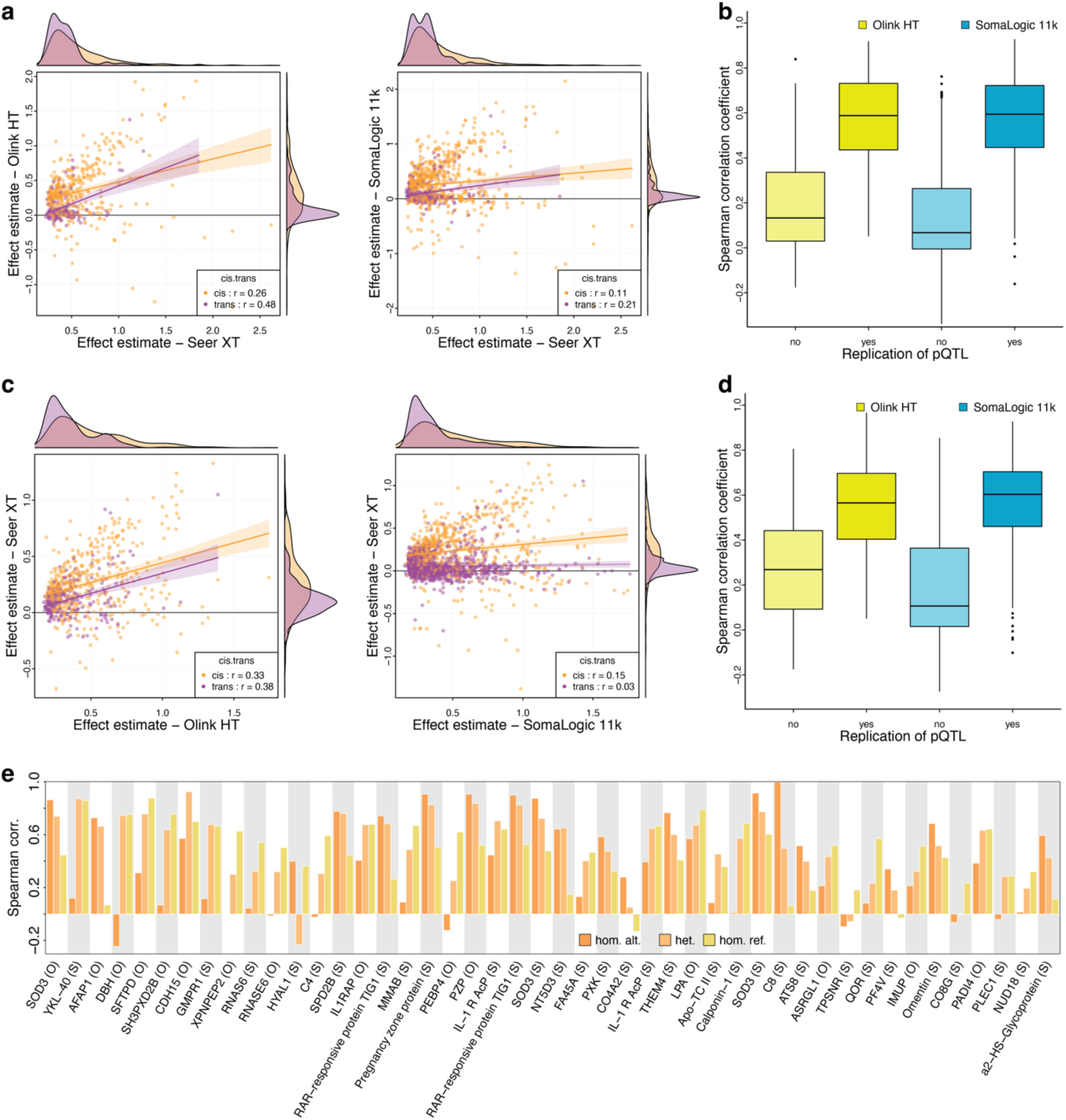
Cross-platform consistency of pQTLs. **a** Effect estimates for 510 (Olink HT) and 1009 (SomaLogic) variant – associations discovered using MS that overlapped with affinity-based measurements. Correlation coefficients are given stratified by cis- and trans-pQTLs. **b** Boxplots indicating the distribution of Spearman correlation coefficients of protein targets stratified by whether or not the associated genetic variant replicated using affinity-based techniques. **c-d** Same as **a** and **b**, but now using 661 (Olink HT) and 2096 (SomaLogic 11k) genetic variant – protein associations discovered using affinity-based techniques and testing for replication using MS-based measurements. **e** 46 variant – protein pairs with robust statistical evidence (see Main text) that the correlation with MS-based measurements differed according to genotype.

### Intersection of pQTLs with the phenome

We next sought to understand whether identified pQTLs have been associated with downstream consequences transcending the proteome and hence point to putative unknown biological mechanisms. For almost half of the pQTLs (n=425), we observed one or more non-proteomic phenotype reported in the GWAS Catalog^38^ for the same variant or their proxies (r^2^>0.6; **Fig. 4a** and **Supplementary Tab. 9**). This included 165 unreported cis-pQTLs, 68 associated with ≥5 traits, that may guide effector gene assignment (**Fig. 4a**). For example, we identified a common variant (chr5:122073485:G>C) intronic of *LOX* associated with decreased plasma levels of the enzyme lysyl oxidase (beta=-0.49; p-value<1.4x10^-38^), the protein product of *LOX*. Lysyl oxidase is secreted by fibrogenic cells and oxidises lysine residues to promote covalent cross-linking of elastin and collagen fibres to stabilize and insolubilize the ECM^39,40^. A genetically mediated lower activity of LOX, as predicted from lower plasma levels, is in line with separate studies linking the same variant to a thicker cornea^41^, taller height^42^, and less youthful appearance^43^. In contrast, the most pleiotropic trans-pQTL or its proxies mapping to *NINJ1* have so far not been reported for any trait providing further evidence for a confined effect on the plasma proteome or a putative role in diseases historically hard to subject to GWAS such as host - pathogen interactions for which a role of *NINJ1* has been suggested based on experimental work^29^.

**Figure 4.**
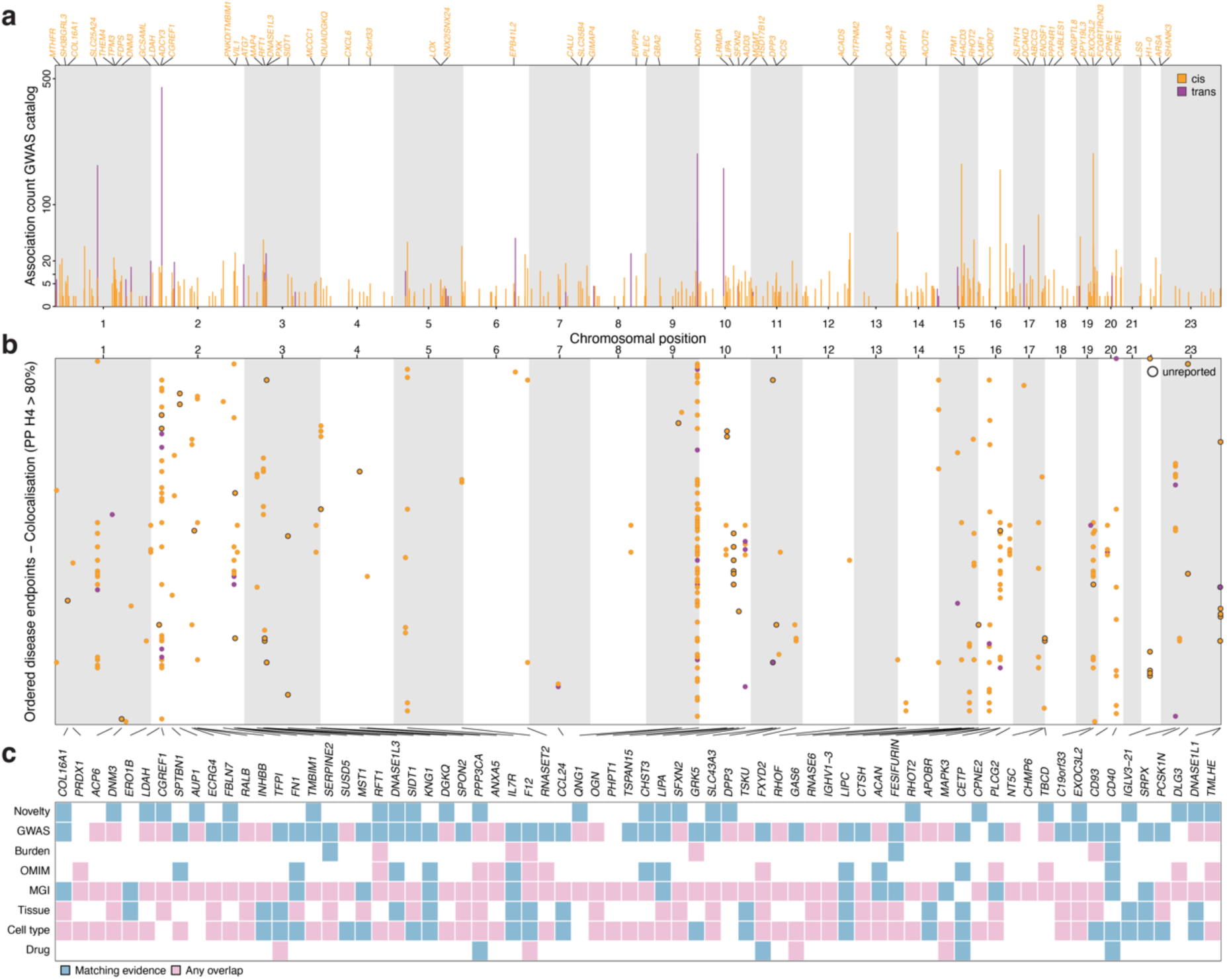
pQTLs link proteins to diseases. **a** Number of associated non-proteomic phenotypes reported in the GWAS catalog for pQTLs or their proxies (r2>0.6; displayed by genomic position) discovered using the Seer Proteograph XT platform. pQTLs mapping to unreported cis-pQTLs and associated with ≥5 non-proteomic outcomes were annotated on top. **b** Results from systematic colocalization analysis of pQTLs with 401 ICD10-coded disease endpoints based on a meta-analysis of FinnGen and UK Biobank44 (y-axis). Only colocalization with strong support of a shared genetic signal between the protein and disease endpoint are shown (posterior probability (PP) of H4 > 80% and respective regional sentinels are in high LD (r2>0.8)). Colocalization with so far unreported pQTLs are highlighted by black circles. **c** Additional layers of evidence that may link the colocalizing protein signal with one or more of the associated diseases. The colour code indicates whether any entry was reported for the protein coding gene in each of the databases and blue indicates direct relevance to the disease colocalizing with the cis-pQTL. Novelty = whether the cis-pQTL has been reported, GWAS = whether any of the colocalizing disease signals has been reported in the GWAS catalog; Burden = matching evidence from a burden of rare predicted loss of function variants45; OMIM = evidence that a rare disorder linked to the protein encoding gene shares phenotypic similarities with any of the colocalizing diseases46; MGI = evidence that any mouse phenotype for the protein encoding genes shares phenotypic similarities with the colocalizing diseases based on the Mouse Genome Informatics initiative47; Tissue/Cell-type = Evidence for enhanced expression of the protein coding gene in disease-relevant tissues and/or cell types48; Drug = Evidence that the protein is the target of an approved drug or a drug in clinical development and the genetic signal is associated with the indication of the drug49

### Multi-level evidence for disease-mediating proteins

To robustly identify a potentially disease mediating role of proteins, we systematically tested whether identified pQTLs also associated with the risk for one or more of 402 diseases in ∼1 million people^44^. We identified a total of 690 pQTL – protein – disease links with strong evidence for a shared genetic signal (posterior probability (PP) > 80%) encompassing 92 unique pQTLs (70 cis-pQTLs) and 136 diseases (**Fig. 4b** and **Supplementary Tab. 14**). While the restricted phenotype and disease overlap may have accounted for fewer associated pQTLs as compared to the intersection with the GWAS Catalog, nine pQTLs (7 cis-pQTL) were exclusively but robustly linked to disease risk in the systematic meta-analysis. This illustrates the need to complement disease-centric efforts, as mostly reported in the GWAS Catalog, with comprehensive biobank screens enabled by electronic health record linkage.

A third of the cis-pQTLs (n=26) mapped to protein encoding genes not previously implicated via proteogenomic evidence. For example, we identified a shared genetic signal between the cis-pQTL chr2:218282080:G>A (p.Pro21Leu; CADD score: 24.2) decreasing plasma levels of transmembrane BAX inhibitor motif-containing 1 (TMBIM1, beta = -0.34; p-value<1.9x10^-19^) and a decreased risk for type 2 diabetes (PP=83.2%; beta = -0,03; p-value<7.0x10^-6^) but an increased risk for cholelithiasis (PP=91.9%; beta = 0.05; p-value<5.6x10^-20^) (**Supplementary Fig. 9**). Our observations align with previous studies at this locus that has been reported for a variety of phenotypes and diseases (n=26) as diverse as blood cell measures, body fat distribution, bone mineral density, colorectal cancer, inflammatory bowel disease, or blood pressure (**Supplementary Fig. 9**). Confident effector gene assignment, however, has been difficult since multiple functional variants for different genes are in high LD, including completely overlapping candidate genes *TMBIM1* and *PNKD* for both of which we observe cis-pQTLs in high LD (r^2^=0.76) (**Supplementary Fig. 9**). The higher posterior probability of a shared signal with the cis-pQTL for TMBIM1 (PP=83.2%) compared to PNKD (PP=50.6%) for type 2 diabetes, even more so for cholelithiasis (91.9% vs 18.4%), aligns with recent evidence that low TMBIM1 by gene knock out can facilitate adipocyte expansion in white adipose tissue rather than adipogenesis, improving obesity-related diseases in mouse models^50^. The latter observation aligns with a decreasing effect of the cis-pQTL on waist-to-hip ratio adjusted for body mass index, a measure of central rather than peripheral fat storage associated with an adverse cardiometabolic health profile^51^.

To overcome ambiguities in effector gene, and hence candidate causal protein assignment, we included additional layers of orthogonal evidence, such as rare gene burden analyses^45^, Mendelian disorders^46^, mouse models^47^, as well as tissue and single cell-type gene expression patterns^48^ (**Fig. 4c**). Among the most compelling examples were plasma levels of CD40 linked to the risk of diverse immune-related diseases, like multiple sclerosis or autoimmune hyperthyroidism, but also non-Hodgkin lymphoma (**Fig. 4b** and **Supplementary Tab. 14**) through different independent layers of evidence. CD40 is a target of high pharmacological interest, with 10 different drugs currently undergoing or having undergone up to phase II trials for 48 indications according to the OpenTargets platform^49^. Importantly, solid organ malignancies are targeted by agonists to enhance the immune system whereas immune-mediated disease, such as psoriasis but also lymphomas, are targeted by inhibitors. These strategies were only partially corroborated by our genetic results indicating opposing effects among immune-system related disorders (**Supplementary** Fig. 10). While positive associations of the CD40 increasing G-allele of chr20:46119308:T>G with Grave’s disease support inhibition^52^ (beta=0.15, p-value<1.1x10^-9^), opposing associations with multiple sclerosis (beta=-0.11; p-value<8.1x10^-6^) and the risk of non-follicular lymphoma (beta=-0.14; p-value<1.9x10^-9^) would rather indicate agonism. Notably, an inverse association with the risk of atrial fibrillation and flutter (beta=-0.04; p-value<4.1x10^-6^) may represent a potential safety signal for CD40 inhibition.

### IgG antibodies carrying IGLV3-21 may predispose to Grave’s disease

We identified notable examples of unreported putative disease mechanisms among the 21 protein – disease(s) connections with evidence from two or more sources (**Fig. 4c**). A common missense variant in *IGLV3-21* (chr22:22713054:G>T; p.Asp68Tyr; AF=70.3%) was associated with lower plasma levels of the protein product immunoglobulin lambda variable 3-21 (IGLV3-21) and decreased risk of autoimmune hyperthyroid disease using different diagnostic criteria or related outcomes, most prominently Grave’s disease (PP=99.6%; beta = -0.19; p<1.9x10^-16^) (**Fig. 5a**). The association was remarkably specific (**Fig. 5b-c**). We observed no evidence that other immunoglobulin genes encoded close by could explain the signal using bulk RNAseq and single cell gene expression data (**Fig. 5b**). We further observed no strong evidence any other disease, including thyroid or autoimmune disease, in our analysis (minimum p-value: 3.6x10^-3^ for chronic lymphoid leukaemia; **Fig. 5c**) but also in large databases^53^ (**Supplementary Tab. 15**). The missense variant overlaps with a complementarity-determining region 2 of IGLV3-21^32^, which, along with other hypervariable regions of Heavy- and Light-chains, determine the binding activity of antibodies to antigens. IGLV3-21 containing antibodies are produced by B-cells^54^ and the same genetic variant associated with lower plasma protein levels specifically associated with lower *IGLV3-21* expression in naïve and memory B-cells^55^ (**Fig. 5a**). Autoantibodies produced by B-cells that activate the thyroid-stimulating hormone receptor (TSHR), called TSH receptor antibodies (TRAb), are the cause of Grave’s disease and lead to secretion of thyroid hormones to the circulation that is not under control of a negative endocrine feedback loop producing a persistent hyperthyroid state^56^ (**Fig. 5d**). Specifically, TRAbs have been shown to be of immunoglobulin class G 1 with lambda light chains^57^, although the variable V3-21 region was not observed in the single patient-derived TRAbs.

Our findings might be of particular importance to guide the development of disease-modifying treatments. Current first line treatment for Grave’s disease includes antithyroid medications, with about 50% of patients showing relapse after 12-18 months ^56^, subsequently necessitating more radical treatment with either radioactive iodine (I^131^) or thyroidectomy. Targeting B-cells is actively pursued, among others, as an alternative strategy to preserve thyroid function in patients with Grave’s disease following the success for the treatment of other autoimmune diseases^58,59^, like multiple sclerosis^60^. We provide human genetic evidence that in contrast to other proposed B-cell related targets, like CD40 (**Supplementary Fig. 10**), specifically targeting B-cells that express IGLV3-21 could provide an alternative route that may minimise adverse effects triggering the onset of other autoimmune disorders as illustrated for CD40. We note that isoforms of IGLV3-21, IGLV3-21*01 and IGLV3-21*02, have so far been only implicated in the development and prognosis of chronic lymphatic leukaemia^61^ and vaccine-induced immune thrombotic thrombocytopenia^62^, respectively.

**Figure 5.**
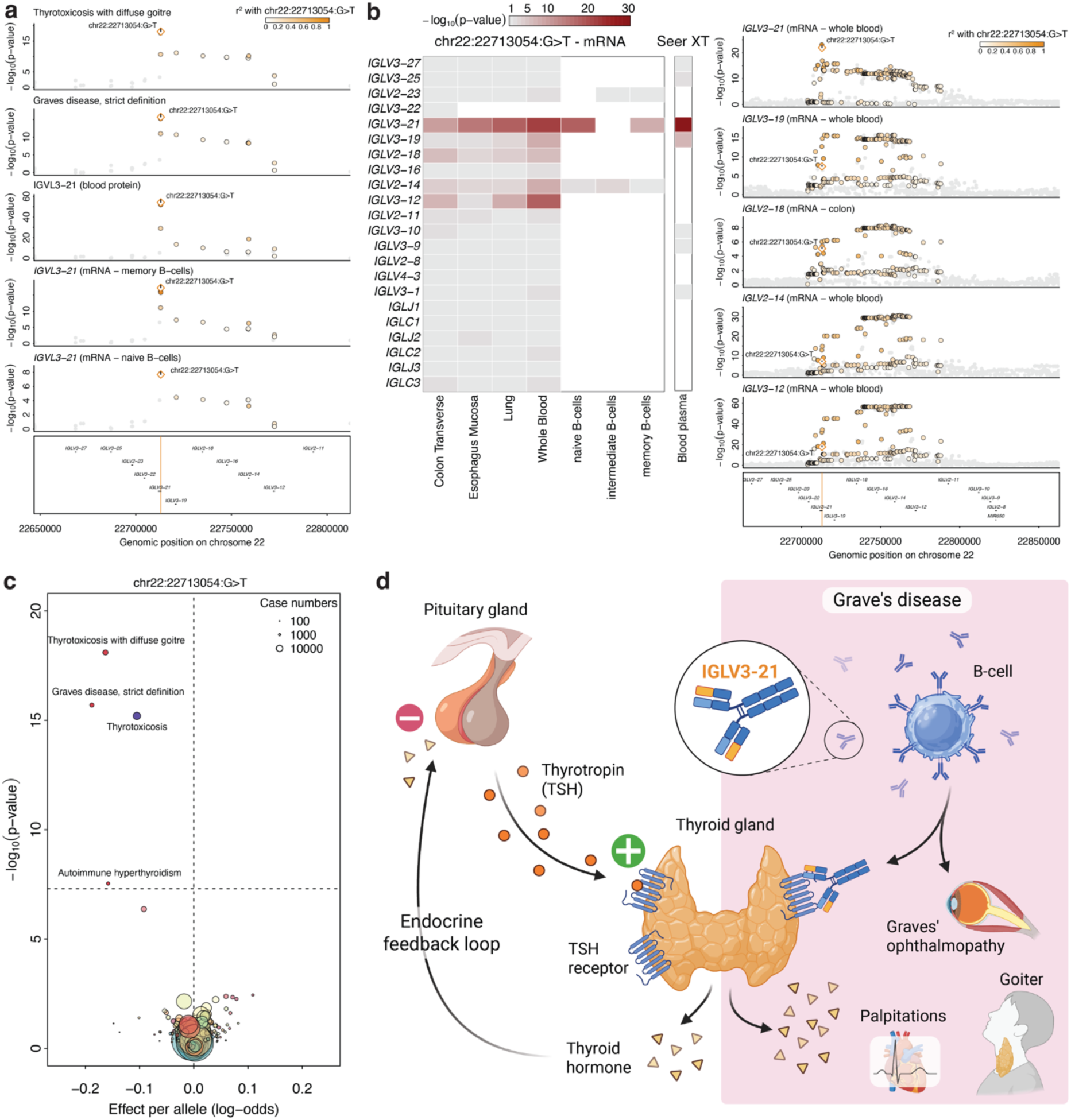
IgG antibodies carrying IGLV3-21 may predispose to Grave’s disease. **a** Stacked regional association plot centred around *IGLV3-21*. Association statistics for thyroid disorders were obtained from a meta-analysis of UK Biobank and FinnGen44. Plasma protein associations from the present study, and mRNA expression from single cell RNA expression of B-cells from the TenK10K consortium55. SNPs (dots) are coloured by LD with the lead cis-pQTL (chr22:22713054:G>T). **b** Heatmap (left) and stacked regional association plot (right) demonstrating specificity of the association of chr22:22713054:G>T with other encoded immunoglobulin genes at the locus across B-cell populations and tissues based on GTEx v863. White fields in the heatmap indicate missing expression of the gene in the respective tissue. **c** Association statistics for chr22:22713054:G>T across 402 diseases based on the same meta-analysis. **d** Schematic of the possible mechanism linking IGLV3-21 expression to Grave’s disease (created with BioRender.com).

## DISCUSSION

Profiling the abundance of thousands of proteins from blood can guide the identification of diverse disease mechanisms, including potential avenues for novel pharmacological interventions, in living humans when successfully linked with germline variation in the genome. Here, we use three complementary plasma proteomic technologies to demonstrate that: 1) untargeted MS-based plasma proteomics covers a unique fraction of the plasma proteome and that poor agreement across platforms is driven by technical factors predicting that ∼40% of affinity-based measures are unlikely to produce concordant results across technologies, 2) pQTL discovery greatly benefits from ancestral and technical diversity discovering >600 pQTLs not reported in studies >30-times larger than ours, and 3) unambiguously linking proteins to disease process requires broad coverage of the proteome and phenome to establish specificity of associations and to maximise sensitivity to minimise false negative results to eventually guide drug target discovery and repurposing.

Affinity-based plasma proteomic platforms now target protein measurement of up to half of the protein coding genome. It is uncertain to what extent many of these proteins circulate in blood at detectable quantities. In the present study using nanoparticle-based enrichment of proteins along with state-of-the art mass spectrometry, we detected >8,000 protein groups in blood at least once, almost doubling what has been summarized recently across >100 studies^20^. One-third of these were not captured by the most comprehensive affinity-based techniques. Even for overlapping protein targets we replicated observations of poor cross-platform concordance^21,26^ that were largely explained by technical factors rather than characteristics of the protein targets. Machine learning models trained on that knowledge indicated the need to improve assay performance across all technologies, with affinity-based assays showing lowest predicted concordance, but higher precision for selected targets compared to the MS-based technology. The role of technical factors contributing to poor cross-platform correlation have been described in detail elsewhere^13–15^, but we add some notable observations. Firstly, CVs were important factors explaining poor correlations across technologies but must be interpreted in relative terms within a technology. That is, a lower CV for a protein in one technology does not necessarily translate into more reliable measurements. Secondly, investigating measurement correlations at the peptide instead of protein group-level showed little evidence for affinity reagents preferring certain protein isoforms that may explain poor correlations and further exemplified the need to improve peptide to protein mappings in MS-based efforts. Thirdly, MS-based plasma proteomics is a relative measure of protein composition (due to injecting a constant amount of protein into the MS) rather than direct protein abundance as inferred by affinity-based reagents, which may have accounted for measurement discrepancies not explained by technical factors. The latter is of particular relevance if plasma samples are enriched for proteins originating from the lysis of blood cells that have been recently shown to strongly determine protein coverage of non-targeted MS-based techniques^64^.

The vast majority of pQTL studies have so far been done in participants of European ancestry^7–11^, but multiple noteworthy examples already demonstrated the value of non-European cohorts even at modest sample size^12,18,65^. The comparatively small yield of ancestral-specific or enriched signals in our study compared to efforts in people of African^12,66,67^ ancestry aligns with our previous work investigating the concordance of common (MAF>1%) genetic variation on laboratory tests in the same population^68^. This is likely a result of being best powered for such common genetic variants that are likely shared across ancestries^69^. However, a unique feature of our study was the enrichment of homozygous carriers of alternative alleles due to a high rate of autozygosity that allowed us to discover non-additive modes of inheritance. This included yet unreported strong autosomal recessive and dominant effects of cis-pQTLs on plasma protein levels with largely undescribed roles in human physiology, such as SLC27A3 or NIPSNAP3 (Nipsnap Homolog 3A), rare genetic variation in an enhancer for the latter was only recently linked to lipid metabolism^70^.

Integrating pQTLs with disease risk loci can guide drug target development or repurposing, but only if the associated protein can be unambiguously assigned to the disease process. An endeavour complicated by the fact, that many genetic variants likely regulate the expression of multiple genes in the proximity^63,71^. A phenomenon we observed here at the protein level as well, but that is this still mostly hidden by the limited coverage and accessibility of proteins in plasma. In other words, even frequently pursued cis-based Mendelian randomization and/or colocalization efforts that work under the, often reasonable, assumption that physical proximity aligns with biological plausibility might still yield false positive results. We therefore collated multiple additional lines of evidence that increase the plausibility of the genetically prioritized link between the protein and disease, including expression in disease-relevant cell-types (like the IGLV3-21 example) or concordant evidence from mouse models (like the TMBIM1 example), but also deprioritized others (like Factor XII and peptic ulcers). However, the scattered evidence across domains illustrated the need to further improve and conceptualize such efforts to confidently link proteins to disease in humans and hence guide the development of new medicines. Such efforts will also benefit from incorporating the full scale of human health and disease associated with specific genetic variation to identify potential treatment options that minimize adverse effects, like we demonstrated for Grave’s disease.

Our study is distinguished by the breadth of proteins covered and depth of genetic profiling in an underrepresented population, but multiple limitations need to be considered when interpreting our results. Firstly, while we applied rigorous correction for multiple testing, replication of results in independent studies, including non-additive effects, are warranted. This includes testing for transferability to other ancestries. Secondly, while nanoparticle enrichment greatly improved coverage of the plasma proteome, it is currently unclear whether this step selectively omits protein targets with unfavourable binding characteristics or misses isoforms that greatly differ in charge from the cognate protein. Thirdly, while our approach to genetically link proteins to diseases distinguished by using separate studies and rigorous methods, mismatches in LD-structure might have led to the underestimation of truly shared signals. Lastly, plasma proteomics provides a readout of protein abundance in blood, which might be a reliable read out for disease relevant mechanisms in cell types or tissues, when combined with human genetic inference, but might miss genetic effects altering protein function but not abundance.

## ACKNOWLEDGEMENTS

Genes & Health is/has recently been core-funded by Wellcome (WT102627, WT210561), the Medical Research Council (UK) (M009017, MR/X009777/1, MR/X009920/1), Higher Education Funding Council for England Catalyst, Barts Charity (845/1796), Health Data Research UK (for London substantive site), and research delivery support from the NHS National Institute for Health Research Clinical Research Network (North Thames). We acknowledge the support of the National Institute for Health and Care Research Barts Biomedical Research Centre (NIHR203330); a delivery partnership of Barts Health NHS Trust, Queen Mary University of London, St George’s University Hospitals NHS Foundation Trust and St George’s University of London

Genes & Health is/has recently been funded by Alnylam Pharmaceuticals, Genomics PLC; and a Life Sciences Industry Consortium of AstraZeneca PLC, Bristol-Myers Squibb Company, GlaxoSmithKline Research and Development Limited, Maze Therapeutics Inc, Merck Sharp & Dohme LLC, Novo Nordisk A/S, Pfizer Inc, Takeda Development Centre Americas Inc.

We thank Social Action for Health, Centre of The Cell, members of our Community Advisory Group, and staff who have recruited and collected data from volunteers. We thank the NIHR National Biosample Centre (UK Biocentre), the Social Genetic & Developmental Psychiatry Centre (King’s College London), Wellcome Sanger Institute, and Broad Institute for sample processing, genotyping, sequencing and variant annotation. This work uses data provided by patients and collected by the NHS as part of their care and support. This research utilised Queen Mary University of London’s Apocrita HPC facility, supported by QMUL Research-IT, http://doi.org/10.5281/zenodo.438045

We thank: Barts Health NHS Trust, NHS Clinical Commissioning Groups (City and Hackney, Waltham Forest, Tower Hamlets, Newham, Redbridge, Havering, Barking and Dagenham), East London NHS Foundation Trust, Bradford Teaching Hospitals NHS Foundation Trust, Public Health England (especially David Wyllie), Discovery Data Service/Endeavour Health Charitable Trust (especially David Stables), Voror Health Technologies Ltd (especially Sophie Don), NHS England (for what was NHS Digital) - for GDPR-compliant data sharing backed by individual written informed consent.

Most of all we thank all of the volunteers participating in Genes & Health.

A favourable ethical opinion for the main Genes & Health research study was granted by NRES Committee London - South East (reference 14/LO/1240) on 16 Sept 2014. Queen Mary University of London is the Sponsor, and Data Controller.

The work was co-funded by the European Union (ERC, GenDrug, 101116072) to M.P.. Views and opinions expressed are however those of the author(s) only and do not necessarily reflect those of the European Union or the European Research Council. Neither the European Union nor the granting authority can be held responsible for them. The funders had no role in study design, data collection and analysis, decision to publish or preparation of the manuscript. This work was supported by the DFG (German Research Foundation) to K.D. (Walter Benjamin Fellowship, Grant Number: 547107463), and the Friede Springer Cardiovascular Prevention

Center at Charité - Universitätsmedizin Berlin, Germany to A.W. We want to acknowledge the participants and investigators of the FinnGen study

## DATA AVAILABILITY

Individual-level data from Genes & Health are available for bona fide researchers on application (https://www.genesandhealth.org/). Genome-wide summary statistics well be released upon publication.

## CODE AVAILABILITY

Associated code will be deposited on GitHub upon final publication.

## AUTHOR CONTRIBUTIONS

Conceptualization: MP, DvH, CL

Laboratory work: KAH

Data curation/Software: MP, JCZ, AW, MK, LK

Formal Analysis: MP, AW, MK, DvH

Methodology: MP, JCZ, AW, MK, DvH, CL

Visualization: MP, CL

Funding acquisition: MP, DvH, CL

Project administration: DvH, SF, CL

Supervision: MP, DvH, CL

Writing – original draft: MP, DvH, CL

Writing – review & editing: all

## METHODS

### Study design

G&H is a cohort of over 70,000 South Asian (Bangladeshi and Pakistani) individuals living in the United Kingdom^23^ (UK). Inclusion criteria included individuals aged 16 and over, from self-reported Bangladeshi and Pakistani backgrounds. Recruitment has been ongoing since 2015. The baseline assessment was performed at recruitment, upon which participants filled in a brief questionnaire, provided a saliva sample for genotyping and sequencing, and consented to being recalled and to longitudinal linkage to electronic health records (EHR) including data from the UK National Health Service (NHS) primary, secondary care, cancer and death registry. Details of the cohort have been previously described. G&H was approved by the London Southeast NRES Committee of the Health Research Authority (14/LO/1240).

The current study comprises a subset of 1464 participants from the G&H cohort that were recalled for additional phenotyping and blood sampling. EDTA-plasma samples were collected and processed within ≤1 hour and subsequently stored at -80°C until further analysis.

### Proteomic Profiling

#### Seer Proteograph

We performed proteomic profiling in 1612 plasma samples from 1464 G&H individuals using the Proteograph XT Assay^41,42^. Proteins were quantitatively captured via nanoparticle (NP)-associated protein coronas, then denatured, reduced, alkylated, and enzymatically digested with Trypsin and LysC. Resulting peptides were purified, quantified, vacuum-dried overnight, and reconstituted at 50 ng/µL.

Peptides (400 ng per injection; 8 µL) underwent Data-Independent Acquisition (DIA) analysis using a Vanquish NEO nanoLC coupled to an Orbitrap Astral mass spectrometer (Thermo Fisher). Peptides were separated using a trap-and-elute configuration (Acclaim PepMap 100 C18 trap column; 50 cm µPAC analytical column) at a flow rate of 1 µL/min over a 22-min gradient from 5–25% solvent B (0.1% FA in ACN), totalling a 33-min run. Mass spectrometry employed MS1 scans (380–980 m/z; Orbitrap detector; resolution 240,000; 0.6 s cycle; 500,000 ion AGC) and 200 fixed-window MS2 DIA scans (150–2000 m/z; 3 Th isolation windows; Astral detector; 25% collision energy; 50,000 ion AGC).

DIA mass spectrometry data were analyzed using the Proteograph™ Analysis Suite’s cloud pipelines^72^, employing the DIA-NN search engine (v1.8.1)^73^ in library-free, match-between-runs mode. MS/MS spectra were matched against an in silico-generated spectral library based on human protein entries (UniProt UP000005640_9606), incorporating cohort-specific protein variants with minor allele count (MAC) ≥5. Search parameters included trypsin digestion (one missed cleavage), N-terminal methionine excision, fixed cysteine carbamidomethylation, peptide length 7–30 amino acids, precursor mass range 300–1800 m/z, fragment ion range 200–1800 m/z, and mass accuracy set to 3 ppm (MS1) and 8 ppm (MS2). Precursor and protein group FDR thresholds were set at 1%. Precursor-level data were annotated as “plus” (all variations retained), “minus” (all variable sites purged), or “reference” (only alternate variants purged), following Suhre et al. 2024^19^. Libraries were filtered at MAC ≥5 and minor allele frequency (MAF) ≥1%. We used the ‘MAC5’ precursor data for all analyses, including for observational correlations with measurements from other platforms. While this led to a drop in precursors supporting protein group assembly, measurements were still highly correlated (r=0.99 across all measurements) indicating a minimal loss in information.

Raw precursor intensities from DIA-NN were normalized using external synthetic peptide standards (PepCal) spiked into samples to account for LC-MS instrument drift. PepCal peptides were quantified via targeted extraction, quality-filtered based on retention time, intensity variation (CV), and completeness, and converted into fold-change relative to median peptide intensities across all runs. Normalization factors were calculated using the median fold-change across high-quality PepCal peptides, further smoothed over every five consecutive runs. Normalized precursor intensities per (biosample, nanoparticle) pair were aggregated into protein-level intensities for each biosample using the MaxLFQ algorithm (*fast_MaxLFQ*, R *iq* package^74^), minimizing variance in protein group intensities. Protein-level intensities were batch-corrected at the Proteograph plate level using *limma*’s *removeBatchEffect* function.

Following batch correction, we identified a total of 8079 protein groups seen in at least one sample, with 5768 protein groups seen in ≥70% of the samples. Based on a blood sample measured repeatedly on each plate, we observed a median coefficient of variation of 22.8% (IQR: 17.8-30.6%) across 7089 protein groups seen at least 5 times. We subsequently performed principal component analysis on the batch corrected aggregated and log2-transformed protein groups that had less than 10% missing values. We identified a total of 67 samples that significantly differed (p-value<3.1x10^-5^) from a multi-dimensional normal distribution based on the first four principal components using the Mahalanobis distance. Those samples were subsequently excluded from downstream analysis and manual inspection confirmed potential measurement issues rather than rare genetic effects as a source of deviation. For samples measured multiple times, we took only the first one measured leaving a total of 1429 samples for downstream analysis.

For genome-wide association testing only, we selected 5768 protein groups with ≤30% missing values and imputed remaining missing values using the R package miceRanger (v.1.4) to enable computational efficient testing. We applied rank-inverse normal transformation to the resulting complete data matrix to minimize the impact of few outlying measurements on association testing.

#### Olink Explore HT

Proteomic profiling with the Olink Explore HT platform was performed in 1641 samples from 1447 individuals. Details of the Olink platform have been previously described in detail^75^. Briefly, Olink relies on proximity extension assays, which targets proteins by pairs of antibodies conjugated to complimentary oligonucleotides. Protein assays are grouped across eight dilutions blocks, with blocks 1 to 4 having a 1:1 dilution (i.e. including low abundant proteins in plasma), block 5 a 1:10 dilution, block 6 a 1:100 dilution, block 7 a 1:1,000 dilution and block 8 a 1:100,000 dilution (i.e. including the most abundant proteins in plasma). Upon binding to their target protein, hybridization between probes enables amplification and subsequent relative quantification through next generation sequencing. Olink’s internal controls involve an incubation (a non-human antigen with matching antibodies), extension (IgG conjugated with a matching oligo pair) and amplification controls (synthetic double stranded DNA). Additional external controls are included in each plate, namely negative, plate (increased to 5 compared to the 3 included in their previous version of the Explore platform) and sample controls. Normalised protein expression (NPX) values are generated by normalisation to the extension control, log2 transformation and further normalisation to the plate controls. Samples are flagged as a FAIL (and no NPX is calculated) if there are < 10k read counts per sample, or if incubation, extension or amplification controls have <150 counts per sample. Blocks are flagged as FAILs (and no NPX values are calculated) if <3 plate controls or <1 negative control pass quality control. We excluded 19 samples due to 1) >50% of protein assays failed or 2) >50% of protein assays with counts below the average count in negative control samples. Based on a blood sample measured repeatedly on each plate, we observed a median coefficient of variation of 30.55% (IQR: 16.35%-46.53%) across all protein targets. *SomaScan 11k v5*

Proteomic profiling with the SomaScan 11k v5 platform was performed in 1751 samples from 1555 individuals. Details of the SomaScan platform have been previously described in detail^76^. Briefly, SomaScan relies on modified DNA-based aptamers that recognise their target protein. Aptamers are grouped across 3 dilutions bins: 20% (1:5), 0.5% (1:200) and 0.005% (1:20,000). SomaLogic’s workflow includes the generation of scaling factors to account for intra- and inter-plate effects, for a hybridization normalisation, median signal normalisation, plate-scale normalisation and interplate calibration steps. Proteins are quantified with relative fluorescence units (RFUs) and finally normalised Adaptive Normalisation by Maximum Likelihood (ANML). SomaLogic does not provide LOD values. We excluded 4 samples that were strong outliers with a median sample RFU more than 3 standard deviation away from the average of the population. Based on a blood sample measured repeatedly on each plate, we observed a median coefficient of variation of 7.71% (IQR: 5.54%-12.28%), across all protein targets.

### Genotyping and imputation

Genotyping was performed using genomic DNA extracted from saliva samples obtained via Oragene saliva sampling kits. Individuals were genotyped on the Illumina GSA v3 chip + extra multi-disease content. Genotype calling and initial genetic data quality control was carried out using Illumina GenomeStudio version 2.0. Briefly, automated clustering using the GenTrain algorithm was performed using 1970 selected very high-quality samples at a subset of high-quality variants (autosomal variants in Hardy–Weinberg equilibrium with GenTrain scores >0.7, reflecting high-confidence clustering). Iterative rounds of manual and automatic reclustering were then performed to identify low-quality variants and samples, ultimately resulting in a dataset with >99% call rate at 637,829 SNPs. This cluster file was then applied to the remaining ∼50,000 samples to call genotypes. Samples were removed if they had lower call rates per sex than in the original batch (<99.2% for females, <99.5% for males). Of the 54,206 genotyped samples, individuals were removed due to a missing NHS number, discordant gender and genetic sex, implausible genetic duplicates (i.e. non-twin duplicates), resulting in a dataset of 51,176 individuals genotyped at 608,329 autosomal SNPs with a genotyping rate of >99.9%.

Following removal of rare variants (MAF < 0.0001), palindromic variants, and indels, genotypes were imputed to the TOPMED-r3 multi-ancestry imputation panel to genome build hg38 using the TOPMED imputation server^77^. Following imputation we performed SNP quality control, filtering to common (MAF > 0.01) biallelic autosomal variants with <10% missingness, imputation quality (INFO score) >0.7 with no significant (p-value<1×10^-15^) deviation from Hardy–Weinberg equilibrium. Variants with duplicate positions were removed. Imputed dosages outside of the ranges 0−0.1, 0.9 −1.1 or 1.9 −2.0 were set to missing. We removed individuals with >10% missing genotypes. Genetic duplicate samples were identified with KING^78^—10 pairs of probable identical twins were identified, and one of each pair removed. Using principal component analysis, we identified and excluded <10 ancestral outliers who did not cluster with South Asian ancestry reference samples from the Human Genome Diversity Project and 1000 Genomes project^69^. Following a clustering-based procedure to estimate categorical ancestry groupings (Bangladeshi or Pakistani), a further 62 participants were excluded due to ambiguous ancestry. We further excluded participants with missing covariate (age and sex) information, and those not included in the proteomic experiment resulting in a final set of 1413 participants for genome-wide association testing.

### Whole Exome Sequencing

The Broad Institute performed ’Standard Germline Exome v6’ using Twist exome capture reagents and Illumina 150bp PE Novaseq 6000 sequencing. BWA-MEM was used to map to the reference genome hg38 with ALT contigs to produce single individual gVCF and cram output files. Preprocessing and variant calling was performed using the Exome Germline Single Sample 3.0.0 pipeline^79^ using Picard 2.23.8^80^, GATK 4.2.2.0 HaplotypeCaller^81^, and Samtools 1.11^82^. The Broad Institute performed basic quality control statistics and delivered crams with >85% bases at >20x Twist bait target coverage. Chromosome Y and MT variants were not called, and chromosome X variants were called diploid for females and males.

Sample quality control (QC) was applied to remove those with <85% bases at >20x coverage in Gencode exons; with the contamination estimate freemix >0.03; self-stated gender that did not match biological sex inferred from exome data (and could not be reconciled); without a valid NHS number, sample duplicates (the lowest coverage sample(s) were removed). Joint genotype calling was performed on these samples using HAIL and GATK GenotypeGVCFs using the Broad Institute Joint Genotyping pipeline^79^. WES crams were compared to 44,396 Illumina GSAv3 chip genotyping samples by using 3,596 common (MAF>0.001) SNPs that are in both WES and GSA hard called genotypes (without imputation) to identify high confidence matches. WES samples with mismatches were removed after comparing them to the GSA data as likely recruitment or laboratory errors. We further removed samples not predicted to be of South Asian ancestry based on PCA and reference samples from the 1000 Genomes Project. Individuals were stratified into Bangladeshi, Pakistani and other South Asian groups based on PCA. Samples were further excluded if the number of SNVs, transition/transversion ratio, number of transitions, number of transversions, number of insertions, number of deletions, insertion/deletion ratio were outside the median ±6 median absolute deviations (MAD) compared to samples from the same population, or if the heterozygote/homozygote ratio, heterozygosity rate was higher than the median + 6 MADs (to avoid removing samples with high autozygosity).

A random forest model was trained on chromosome 20 using the true positive (high confidence variant sites discovered in the 1,000 Genomes Project, SNVs present on the Illumina Omni 2.5 genotyping array and found in the 1,000 Genomes Project^69^, INDELs present in the Mills and Devine data^83^, HapMap3^84^ SNVs and INDELs) and false positive variants (QD<2 OR FS>60 OR MQ<30) as described above and then applied to the whole dataset. Features selected included quality by depth (QD), mean heterozygous allele balance, multiallelic site, strand odds ratio (SOR), mapping quality (MQ), number of alternative alleles at a site, variant type, site for which alleles include a ‘*’ allele, rank sum test for mapping qualities of reference versus alternative reads, multiallelic site containing SNVs and indels, rank sum test for relative positioning of reference versus alternative alleles within reads and allele type. Variants were ranked by their random forest score (i.e., the probability a variant reflects a true positive) and binned. We selected a random forest bin of 77 (true positive rate = 98.76%, false negative rate = 0.70%) and 57 (true positive rate = 95.65%, false negative rate = 4.88 %) to retain SNVs and indels, respectively.

Among variants passing variant QC criteria, we further tested a combination of random forest bin, depth (DP), genotype quality (GQ), heterozygous allele balance (hetAB) and call rate to perform genotype-level QC. Specifically for variants passing the given random forest bin, genotypes were set to missing if they did not pass one or more of the listed genotype QC criteria. To determine the optimal combination, we calculated the percentage of true and false positives, the transmitted / untransmitted synonymous singleton ration in trios (inferred using KING^78^), the total number of Mendelian errors in trios and the mean number of heterozygous calls in runs of homozygosity (ROHs), for each combination of QC filters. For X chromosome variants, DP and GQ thresholds were tested separately for males and females and we calculated the mean number of heterozygous calls in non-pseudoautosomal regions in males instead of number of heterozygous calls in ROHs. The final set of filters selected were random forest bin 80, DP 10, GQ 20, hetAB 0.2 for SNPs, and random forest bin 75, DP 10, GQ 20, hetAB 0.2 and call rate 0.95 for SNVs; and random forest bin 40, DP 10, GQ 20, hetAB 0.3 and call rate 0.95 for indels. For subsequent genetic analyses, we retained 1418 participants with available MS-based proteomic data that passed quality control.

### Cross-platform protein target mapping and annotation

We extracted from each vendor UniProt identifiers^85^ for protein groups (MS) or affinity targets that resulted in a list of 14,757 unique identifiers (note that some MS-based protein groups map to multiple UniProt identifiers). We used UniProt identifier omitting isoform information that was only available for MS-based assays. For each unique UniProt identifier, we then mapped all platform-specific IDs that mentioned in their description the respective UniProt identifier to identify unique and overlapping targets across proteomic platforms. We subsequently used different databases^48,85–87^ to annotate protein coding genes, their genomic position (GRCh build 38), different protein characteristics, tissue- and cell-type expression patterns of protein coding genes, as well as predictions of secretome locations. Missing information on the genomic location of protein coding genes was manually obtained from GeneCards^88^.

### Statistical analysis

We computed missingness rates for protein targets across platforms based on missing peptide information using MS-based assays, or by computing a pseudo lower limit of detection for affinity-based assays by taking the median signal for buffer samples and adding 4 times the median absolute deviance measured across repeated samples. We computed coefficient of variations and associated measures based on a biological replicate measured on each plate across proteomic technologies. We further computed standard distribution measures such as mean, standard deviation, or median as well as tested for multi-modal distributions using a dip test.

As an indicator of measurement concordance based on semi-quantitative assay readouts, we computed Spearman correlation coefficients for all pairwise overlapping targets among 1396 unique participant samples who had measurements available on all three platforms. For affinity-based targets, we additionally tested whether stratifying computation of Spearman correlation coefficients by genotypes of associated cis-pQTLs improved concordance with MS-based measurements by 1) testing for an interaction effect of the SNP with MS-based measurements, and 2) computing p-values for differences in correlation coefficients as implemented in the R package *cocor* (v.1.1.4)^89^.

For each of the three possible pairwise-platform comparisons, we used Boruta feature selection^90^ (R package Boruta v.9.0) to identify features significantly (p<0.05 after multiple testing) associated with the gradient of correlation coefficients across overlapping protein targets. To this end, we computed platform-specific quality control measures (**Supplementary Tab. 8**) but also integrated information on genetic findings (see below) and protein characteristics. To estimate the predictive performance of machine learning models, we trained a Random Forest model to predict Spearman correlation coefficients using 10-fold cross validation and parameter tuning as implemented in the *caret* and *RandomForest* R package (V7.0 and V4.7). Having successfully trained models for each pairwise-platform comparison, we trained a final model for each platform using the best Spearman correlation value for each overlapping protein target irrespective of the proteomic platform. We only utilised QC-measures specific to the platform of interest as well as generic protein characteristics as features. The final set of models for each platform, again trained using 10-cv, was then used to predict Spearman correlation coefficients even for proteins unique to the respective platform. We computed the median of predicted correlation coefficients across all 10 models per platform as a final measure. We used similar feature prioritization and prediction frameworks to test for the replication of pQTLs, either any or specifically those close to the protein coding gene. For the latter, we added bespoke variant and gene characteristics to the feature set derived from the variant effect predictor tool (VEP v111), including annotation of protein altering variants or and changes in amino acid composition on protein characteristics.

### Genome-wide association analysis

We run genome-wide association testing for plasma levels of each of 5678 protein groups using the REGENIE software^91^ (v.4.0). For step 1, we used an LD-pruned (r^2^<0.2 or <0.9, respectively) set of common (MAF>1%) genotyped (GWAS) or sequenced (WES) markers with high coverage (missingness < 1%). We included sex, age, inferred ancestry (Bangladeshi or Pakistani ancestry), and the first twenty genetic principal components as covariates. In step 2, we run association testing again separately for imputed and sequenced genetic variants using the same set of covariates and default parameters for REGENIE.

### Integrated statistical fine-mapping and novelty assignment

To integrate results from GWAS and ExWAS, we first extracted regional lead signals (p<8.6x10^-12^ correcting genome-wide significance by 5768 tested protein groups) in a 3Mb window from each file set separately. After collating associated regions across GWAS and ExWAS results files, we used statistical fine-mapping^92^ as implemented in the R package *susieR* (v.0.14.2) to identify independent credible sets (maximum r^2^<0.25). For each region, we retained exonic variant based on exome sequencing or imputed genetic variants otherwise. We sequentially tested for the presence of 1 to at most 10 credible sets, and afterwards computed for each iteration the LD among all possible lead credible set variants. We retained the maximum number of credible sets that still delivered approximately independent credible sets. We used combined genetic information on 690 unrelated participants of the Genes & Health cohort with available proteomic data to compute the LD-backbone for fine-mapping. We finally filtered fine-mapping results to only retain credible sets for which the lead variant passed corrected statistical significance. This integrated approach allowed us to identify independent coding and non-coding pQTLs in the same region without additional computational burden of running conditional analysis.

For each identified lead credible set variant – protein group pairing, we intersected the associated region with ones reported in 17 previous large-scale proteomic studies up to 30 larger than ours^4–8,11–14,16,17,37,93–97^. If the region was not reported, we classified the paring as ‘novel locus’. In case the region was reported, but for a different set of protein targets (based on UniProt), we classified the association as ‘novel protein at locus’ distinguishing between whether or not the protein has been targeted by previous studies. For lead credible set variants mapping to an existing locus – protein associations, we further computed the LD-backbone in G&H and asked whether any of the variants in at least weak LD (r^2^>0.1) contained a reported signal and flagged the association accordingly. To finally flag potential ancestral-specific or enriched signals, we compared allele frequency in the G&H cohort with those reported in gnomAD for Non-finish Europeans and flagged those with strong evidence of being rare in the latter.

### Testing for non-additive effects

For each lead credible set variant – protein pair, we tested for a potential departure from an additive genetic model by re-running association testing for this pair but introducing another term encoding heterozygous carriers into the model following previous work^98^. Briefly, the additional term is a generic term to test for a significant departure from linearity across the three genotypes and we subsequently assigned potential models of inheritance based on computing log-likelihood ratio tests recoding the genotype according to three different models of inheritance based on the annotated reference allele. To distinguish between truly recessive/dominant effects and other deviations from normality, we additionally computed the gain in significance of either the dominant or additive model as suggested previously and kept only variant – protein association examples with a gain>1^99^. These analyses were implemented in R v.4.3.1 using the REGNIE step 1 files to account for relatedness.

### Effector gene assignment

For each lead credible set variant and proxies in strong LD, we queried multiple data bases to facilitate effector gene assignment. Those included data from the ABC CATlas^100^, fine mapped eQTL catalogue data^101^, evidence from HiC experiments^102,103^, OMIM^46^, or OpenTargets^49^.

To rank candidate effector genes within a locus of interest, we designed a pragmatic but transparent scoring system. We assigned points of 1 if a gene in the locus was linked to the variant if 1) it was the closest gene, 2), overlapping an exon, 3) the protein target encoding gene (‘cis’), 4) encoded a protein complex or ligand receptor partner (‘trans’), 5) there was eQTL evidence (posterior inclusion probability (PIP) > 0.1), 6) there was evidence from ABC score, 7) evidence that the section of the genome was implicated by Hi-C to the expression of the gene in a tissue or cell-type.

### Phenotypic follow-up of pQTLs

To assess, whether pQTLs, ie., genetic signals merged across all protein targets, might have been already reported for non-proteomic traits, we computed LD-proxies (r^2^>0.6) for each lead credible set variant and intersected those with the GWAS catalog (download: 03/02/2025). For each unique trait, we only kept the genetic variant in highest LD or the pQTL itself to avoid double counting. Before doing so, we filtered the GWAS catalog results for signals meeting at least genome-wide significance, reporting of risk alleles, and omitting blood proteomic studies.

We systematically tested for a potentially shared genetic architecture between pQTLs and 402 diseases based on a meta-analysis of FinnGen and UK Biobank provided by the FinnGen consortium. We restricted our search to endpoints with 3 or more genome-wide association signals to ensure enough power to test for colocalization. To minimize computational burden, we first queried all pQTLs or proxies thereof in GWAS summary statistics across all 402 endpoints and did only put those regions forward for colocalization with evidence for a potentially significant effect (p<10^-6^ accounting for the number of signals and outcomes tested). We used default priors for colocalization apart from p_12_ which we set to 5.0x10^-6^ as recommended previously^104^. We used colocalization under the one causal variant assumption and further computed LD between regional lead variants to filter potential false-positive colocalization signals (r^2^<0.8). We opted for such a conservative strategy due to non-matching LD backbones across studies and acknowledge that this strategy might have missed true positive results reported elsewhere.

## REFERENCES

1. Suhre, K., McCarthy, M. I. & Schwenk, J. M. Genetics meets proteomics: perspectives for large population-based studies. Nat. Rev. Genet. 22, 19–37 (2021).

2. Zheng, J. et al. Phenome-wide Mendelian randomization mapping the influence of the plasma proteome on complex diseases. Nat. Genet. 52, 1122–1131 (2020).

3. Sun, B. B. et al. Plasma proteomic associations with genetics and health in the UK Biobank. Nature (2023) doi:10.1038/s41586-023-06592-6.

4. Dhindsa, R. S. et al. Rare variant associations with plasma protein levels in the UK Biobank. Nature 622, 339–347 (2023).

5. Folkersen, L. et al. Genomic and drug target evaluation of 90 cardiovascular proteins in 30,931 individuals. Nat. Metab. 2, 1135–1148 (2020).

6. Koprulu, M. et al. Proteogenomic links to human metabolic diseases. Nat. Metab. 5, 516–528 (2023).

7. Suhre, K. et al. Connecting genetic risk to disease end points through the human blood plasma proteome. Nat. Commun. 8, (2017).

8. Sun, B. B. et al. Genomic atlas of the human plasma proteome. Nature 558, 73–79 (2018).

9. Emilsson, V. et al. Co-regulatory networks of human serum proteins link genetics to disease. Science 361, 769–773 (2018).

10. Pietzner, M. et al. Mapping the proteo-genomic convergence of human diseases. Science 374, eabj1541 (2021).

11. Ferkingstad, E. et al. Large-scale integration of the plasma proteome with genetics and disease. Nat. Genet. (2021) doi:10.1038/s41588-021-00978-w.

12. Zhang, J. et al. Plasma proteome analyses in individuals of European and African ancestry identify cis-pQTLs and models for proteome-wide association studies. Nat. Genet. 54, 593–602 (2022).

13. Sveinbjornsson, G., Magnusson, M. I., Helgason, A. & Oddsson, A. Large-scale plasma proteomics comparisons through genetics and disease associations. (2023) doi:10.1038/s41586-023-06563-x.

14. Pietzner, M. et al. Synergistic insights into human health from aptamer- and antibody-based proteomic profiling. Nat. Commun. 12, 6822 (2021).

15. Katz, D. H. et al. Proteomic profiling platforms head to head: Leveraging genetics and clinical traits to compare aptamer- and antibody-based methods. Sci. Adv. 8, 5164 (2022).

16. Yao, C. et al. Genome-wide mapping of plasma protein QTLs identifies putatively causal genes and pathways for cardiovascular disease. Nat. Commun. 9, 3268 (2018).

17. Niu, L. et al. Plasma proteome variation and its genetic determinants in children and adolescents. Nat. Genet. 1–12 (2025) doi:10.1038/s41588-025-02089-2.

18. Xu, F. et al. Genome-wide genotype-serum proteome mapping provides insights into the cross-ancestry differences in cardiometabolic disease susceptibility. Nat. Commun. 14, 896 (2023).

19. Suhre, K. et al. Nanoparticle enrichment mass-spectrometry proteomics identifies protein-altering variants for precise pQTL mapping. Nat. Commun. 15, 989 (2024).

20. Geyer, P. E. et al. The Circulating Proteome─Technological Developments, Current Challenges, and Future Trends. J. Proteome Res. 23, 5279–5295 (2024).

21. Beimers, W. F., Overmyer, K. A., Sinitcyn, P., Lancaster, N. M. & Coon, J. J. A Technical Evaluation of Plasma Proteomics Technologies. 2025.01.08.632035 Preprint at 10.1101/2025.01.08.632035 (2025).

22. Proteomics, Q. N. et al. Protein Coronas on Functionalized Nanoparticles Enable Quantitative and Precise Large-Scale Deep Plasma Proteomics. (2023).

23. Finer, S. et al. Cohort Profile: East London Genes & Health (ELGH), a community-based population genomics and health study in British Bangladeshi and British Pakistani people. Int. J. Epidemiol. 49, 20–21i (2020).

24. Malawsky, D. S. et al. Influence of autozygosity on common disease risk across the phenotypic spectrum. Cell 186, 4514–4527.e14 (2023).

25. Uhlén, M. et al. The human secretome. 0274, 1–9 (2019).

26. Kirsher, D. Y. et al. The Current Landscape of Plasma Proteomics: Technical Advances, Biological Insights, and Biomarker Discovery. 2025.02.14.638375 Preprint at 10.1101/2025.02.14.638375 (2025).

27. Chen, X. et al. An autoimmune disease variant of IgG1 modulates B cell activation and differentiation. Science 362, 700–705 (2018).

28. Degen, M. et al. Structural basis of NINJ1-mediated plasma membrane rupture in cell death. Nature 618, 1065–1071 (2023).

29. Kayagaki, N. et al. NINJ1 mediates plasma membrane rupture during lytic cell death. Nature 591, 131–136 (2021).

30. Janes, J. et al. Predicted mechanistic impacts of human protein missense variants. 2024.05.29.596373 Preprint at 10.1101/2024.05.29.596373 (2024).

31. Lacoste, J. et al. Pervasive mislocalization of pathogenic coding variants underlying human disorders. Cell 187, 6725–6741.e13 (2024).

32. Stephenson, J. D. et al. ProtVar: mapping and contextualizing human missense variation. Nucleic Acids Res. 52, W140–W147 (2024).

33. Kerimov, N., Hayhurst, J. D., Manning, J. R. & Walter, P. eQTL Catalogue : a compendium of uniformly processed human gene expression and splicing QTLs. 1–34 (2020).

34. Pei, Z. et al. Mouse Very Long-chain Acyl-CoA Synthetase 3/Fatty Acid Transport Protein 3 Catalyzes Fatty Acid Activation but Not Fatty Acid Transport in MA-10 Cells *. J. Biol. Chem. 279, 54454–54462 (2004).

35. Backman, J. D. et al. Exome sequencing and analysis of 454,787 UK Biobank participants. Nature 1–20 (2021) doi:10.1038/s41586-021-04103-z.

36. Raffield, L. M. et al. Comparison of Proteomic Assessment Methods in Multiple Cohort Studies. Proteomics 20, e1900278 (2020).

37. Suhre, K. et al. A genome-wide association study of mass spectrometry proteomics using the Seer Proteograph platform. 2024.05.27.596028 Preprint at 10.1101/2024.05.27.596028 (2024).

38. Buniello, A. et al. The NHGRI-EBI GWAS Catalog of published genome-wide association studies, targeted arrays and summary statistics 2019. Nucleic Acids Res. 47, D1005– D1012 (2019).

39. Mäki, J. M. et al. Lysyl Oxidase Is Essential for Normal Development and Function of the Respiratory System and for the Integrity of Elastic and Collagen Fibers in Various Tissues. Am. J. Pathol. 167, 927–936 (2005).

40. Kagan, H. M. & Li, W. Lysyl oxidase: Properties, specificity, and biological roles inside and outside of the cell. J. Cell. Biochem. 88, 660–672 (2003).

41. Gao, X. R., Huang, H., Nannini, D. R., Fan, F. & Kim, H. Genome-wide association analyses identify new loci influencing intraocular pressure. Hum. Mol. Genet. 27, 2205–2213 (2018).

42. Yengo, L. et al. A saturated map of common genetic variants associated with human height. Nature 610, 704–712 (2022).

43. Roberts, V., Main, B., Timpson, N. J. & Haworth, S. Genome-Wide Association Study Identifies Genetic Associations with Perceived Age. J. Invest. Dermatol. 140, 2380–2385 (2020).

44. Kurki, M. I. et al. FinnGen provides genetic insights from a well-phenotyped isolated population. Nature 613, 508–518 (2023).

45. Jurgens, S. J. et al. Rare coding variant analysis for human diseases across biobanks and ancestries. Nat. Genet. 56, 1811–1820 (2024).

46. Amberger, J. S. & Hamosh, A. Searching Online Mendelian Inheritance in Man (OMIM): A Knowledgebase of Human Genes and Genetic Phenotypes. Curr. Protoc. Bioinforma. 58, 1.2.1–1.2.12 (2017).

47. Bult, C. J. et al. Mouse Genome Database (MGD) 2019. Nucleic Acids Res. 47, D801–D806 (2019).

48. Digre, A. The human protein atlas — Integrated omics for single cell mapping of the human proteome. 1–11 (2023) doi:10.1002/pro.4562.

49. Ghoussaini, M. et al. Open Targets Genetics: systematic identification of trait-associated genes using large-scale genetics and functional genomics. Nucleic Acids Res. 49, D1311– D1320 (2021).

50. Zhao, G.-N. et al. TMBIM1 is an inhibitor of adipogenesis and its depletion promotes adipocyte hyperplasia and improves obesity-related metabolic disease. Cell Metab. 33, 1640–1654.e8 (2021).

51. Lotta, L. A. et al. Association of Genetic Variants Related to Gluteofemoral vs Abdominal Fat Distribution with Type 2 Diabetes, Coronary Disease, and Cardiovascular Risk Factors. JAMA - J. Am. Med. Assoc. 320, 2553–2563 (2018).

52. Kahaly, G. J. et al. A Novel Anti-CD40 Monoclonal Antibody, Iscalimab, for Control of Graves Hyperthyroidism—A Proof-of-Concept Trial. J. Clin. Endocrinol. Metab. 105, 696– 704 (2020).

53. Costanzo, M. C. et al. Cardiovascular Disease Knowledge Portal: A Community Resource for Cardiovascular Disease Research. Circ. Genomic Precis. Med. 16, e004181 (2023).

54. Foreman, A. L., Van de Water, J., Gougeon, M.-L. & Gershwin, M. E. B cells in autoimmune diseases: Insights from analyses of immunoglobulin variable (Ig V) gene usage. Autoimmun. Rev. 6, 387–401 (2007).

55. Cuomo, A. S. E. et al. Impact of Rare and Common Genetic Variation on Cell Type-Specific Gene Expression. 2025.03.20.25324352 Preprint at 10.1101/2025.03.20.25324352 (2025).

56. Wiersinga, W. M., Poppe, K. G. & Effraimidis, G. Hyperthyroidism: aetiology, pathogenesis, diagnosis, management, complications, and prognosis. Lancet Diabetes Endocrinol. 11, 282–298 (2023).

57. Sanders, J. et al. Human monoclonal thyroid stimulating autoantibody. The Lancet 362, 126–128 (2003).

58. Lee, D. S. W., Rojas, O. L. & Gommerman, J. L. B cell depletion therapies in autoimmune disease: advances and mechanistic insights. Nat. Rev. Drug Discov. 20, 179–199 (2021).

59. Schett, G., Nagy, G., Krönke, G. & Mielenz, D. B-cell depletion in autoimmune diseases. Ann. Rheum. Dis. 83, 1409–1420 (2024).

60. Lane, L. C., Cheetham, T. D., Perros, P. & Pearce, S. H. S. New Therapeutic Horizons for Graves’ Hyperthyroidism. Endocr. Rev. 41, 873–884 (2020).

61. Maity, P. C. et al. IGLV3-21*01 is an inherited risk factor for CLL through the acquisition of a single-point mutation enabling autonomous BCR signaling. Proc. Natl. Acad. Sci. 117, 4320–4327 (2020).

62. Wang, J. J. et al. Vaccine-induced immune thrombotic thrombocytopenia is mediated by a stereotyped clonotypic antibody. Blood 140, 1738–1742 (2022).

63. GTEx Consortium. The GTEx Consortium atlas of genetic regulatory effects across human tissues. Science 369, 1318–1330 (2020).

64. Korff, K. et al. Pre-Analytical Drivers of Bias in Bead-Enriched Plasma Proteomics. 2025.05.07.652495 Preprint at 10.1101/2025.05.07.652495 (2025).

65. Wang, Q. S. et al. Statistically and functionally fine-mapped blood eQTLs and pQTLs from 1,405 humans reveal distinct regulation patterns and disease relevance. Nat. Genet. 56, 2054–2067 (2024).

66. Tahir, U. A. et al. Proteogenomic analysis integrated with electronic health records data reveals disease-associated variants in Black Americans. J. Clin. Invest. 134, (2024).

67. Soremekun, O. et al. Linking the plasma proteome to genetics in individuals from continental Africa provides insights into type 2 diabetes pathogenesis. 2024.09.16.24313728 Preprint at 10.1101/2024.09.16.24313728 (2024).

68. Jacobs, B. M. et al. Genetic architecture of routinely acquired blood tests in a British South Asian cohort. Nat. Commun. 15, 8929 (2024).

69. 1000 Genomes Project Consortium et al. A global reference for human genetic variation. Nature 526, 68–74 (2015).

70. Li, X. et al. A statistical framework for powerful multi-trait rare variant analysis in large-scale whole-genome sequencing studies. 2023.10.30.564764 Preprint at 10.1101/2023.10.30.564764 (2023).

71. Tong, P., Monahan, J. & Prendergast, J. G. D. Shared regulatory sites are abundant in the human genome and shed light on genome evolution and disease pleiotropy. PLOS Genet. 13, e1006673 (2017).

72. Guturu, H. et al. Cloud-Enabled Scalable Analysis of Large Proteomics Cohorts. J. Proteome Res. 24, 1462–1469 (2025).

73. Demichev, V., Messner, C. B., Vernardis, S. I., Lilley, K. S. & Ralser, M. DIA-NN: neural networks and interference correction enable deep proteome coverage in high throughput. Nat. Methods (2019) doi:10.1038/s41592-019-0638-x.

74. Pham, T. V., Henneman, A. A. & Jimenez, C. R. iq: an R package to estimate relative protein abundances from ion quantification in DIA-MS-based proteomics. Bioinformatics 36, 2611–2613 (2020).

75. Wik, L. et al. Proximity Extension Assay in Combination with Next-Generation Sequencing for High-throughput Proteome-wide Analysis. Mol. Cell. Proteomics 20, (2021).

76. Gold, L. et al. Aptamer-based multiplexed proteomic technology for biomarker discovery. PLoS ONE 5, (2010).

77. Taliun, D. et al. Sequencing of 53,831 diverse genomes from the NHLBI TOPMed Program. (2021) doi:10.1038/s41586-021-03205-y.

78. Manichaikul, A. et al. Robust relationship inference in genome-wide association studies. Bioinformatics 26, 2867–2873 (2010).

79. Pipelines, W. et al. WDL Analysis Research Pipelines: Cloud-Optimized Workflows for Biological Data Processing and Reproducible Analysis. Preprint at 10.20944/preprints202401.2131.v1 (2024).

80. Releases · broadinstitute/picard. *GitHub* https://github.com/broadinstitute/picard/releases.

81. Poplin, R., et al. Scaling accurate genetic variant discovery to tens of thousands of samples. 201178 Preprint at 10.1101/201178 (2018).

82. Danecek, P. et al. Twelve years of SAMtools and BCFtools. GigaScience 10, giab008 (2021).

83. Mills, R. E. et al. An initial map of insertion and deletion (INDEL) variation in the human genome. Genome Res. 16, 1182–1190 (2006).

84. Altshuler, D. M. et al. Integrating common and rare genetic variation in diverse human populations. Nature 467, 52–58 (2010).

85. Bateman, A. et al. UniProt: The universal protein knowledgebase. Nucleic Acids Res. 45, D158–D169 (2017).

86. Smedley, D. et al. The BioMart community portal: an innovative alternative to large, centralized data repositories. Nucleic Acids Res. 43, W589–W598 (2015).

87. Voisinne, G. queryup: Query the ‘UniProtKB’ REST API. (2023).

88. Stelzer, G. et al. The GeneCards Suite: From Gene Data Mining to Disease Genome Sequence Analyses. Curr. Protoc. Bioinforma. 54, 1.30.1–1.30.33 (2016).

89. Diedenhofen, B. & Musch, J. cocor: A Comprehensive Solution for the Statistical Comparison of Correlations. PLOS ONE 10, e0121945 (2015).

90. Kursa, M. B. & Rudnicki, W. R. Feature selection with the boruta package. J. Stat. Softw. 36, 1–13 (2010).

91. Mbatchou, J. et al. Computationally efficient whole-genome regression for quantitative and binary traits. Nat. Genet. 53, 1097–1103 (2021).

92. Zou, Y., Carbonetto, P., Wang, G. & Stephens, M. Fine-mapping from summary data with the “ Sum of Single Effects ” model. 1–33 (2021).

93. Gilly, A. et al. Whole-genome sequencing analysis of the cardiometabolic proteome. Nat. Commun. 11, 6336 (2020).

94. Gudjonsson, A. et al. A genome-wide association study of serum proteins reveals shared loci with common diseases. Nat. Commun. 13, 480 (2022).

95. Katz, D. H. et al. Whole Genome Sequence Analysis of the Plasma Proteome in Black Adults Provides Novel Insights Into Cardiovascular Disease. Circulation 145, 357–370 (2022).

96. Png, G. et al. Mapping the serum proteome to neurological diseases using whole genome sequencing. Nat. Commun. 12, 7042 (2021).

97. Repetto, L. et al. The genetic landscape of neuro-related proteins in human plasma. *Nat*. Hum. Behav. 8, 2222–2234 (2024).

98. Lotta, L. A. et al. A cross-platform approach identifies genetic regulators of human metabolism and health. Nat. Genet. 53, 54–64 (2021).

99. Scherer, N., Sekula, P., Pfaffelhuber, P. & Schlosser, P. pgainsim: an R-package to assess the mode of inheritance for quantitative trait loci in GWAS. Bioinformatics 37, 3061– 3063 (2021).

100. Zhang, K. et al. A single-cell atlas of chromatin accessibility in the human genome. Cell 184, 5985–6001.e19 (2021).

101. Kerimov, N. et al. eQTL Catalogue 2023: New datasets, X chromosome QTLs, and improved detection and visualisation of transcript-level QTLs. PLOS Genet. 19, e1010932 (2023).

102. Jung, I. et al. A compendium of promoter-centered long-range chromatin interactions in the human genome. Nat. Genet. 51, 1442–1449 (2019).

103. Javierre, B. M. et al. Lineage-Specific Genome Architecture Links Enhancers and Non-coding Disease Variants to Target Gene Promoters. Cell 167, 1369–1384.e19 (2016).

104. Wallace, C. Eliciting priors and relaxing the single causal variant assumption in colocalisation analyses. PLOS Genet. 16, e1008720 (2020).

