## Supplementary Figure for "Nanoparticle enriched mass spectrometry proteomics in British South Asians identifies novel variant-protein-disease mechanisms"

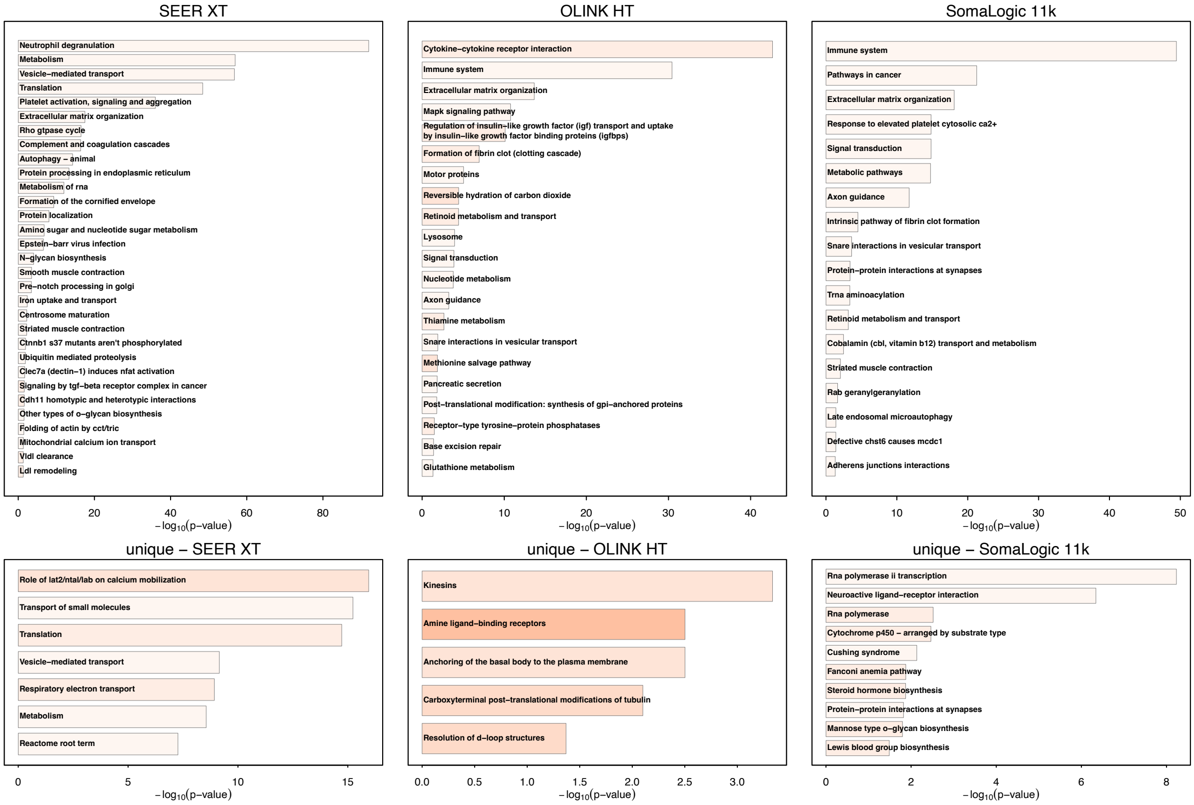


**Supplementary Fig. 1** Enriched pathways among proteins covered by each of the three platforms listed. We reduced the number of redundant pathways by selecting only those that share minimal overlap with the most significant ones. Colour intensity is proportional to the fold enrichment of pathways. The upper panel shows pathways enriched among all proteins covered, whereas the lower panel only shows pathways enriched among protein targets uniquely covered by the platform listed.

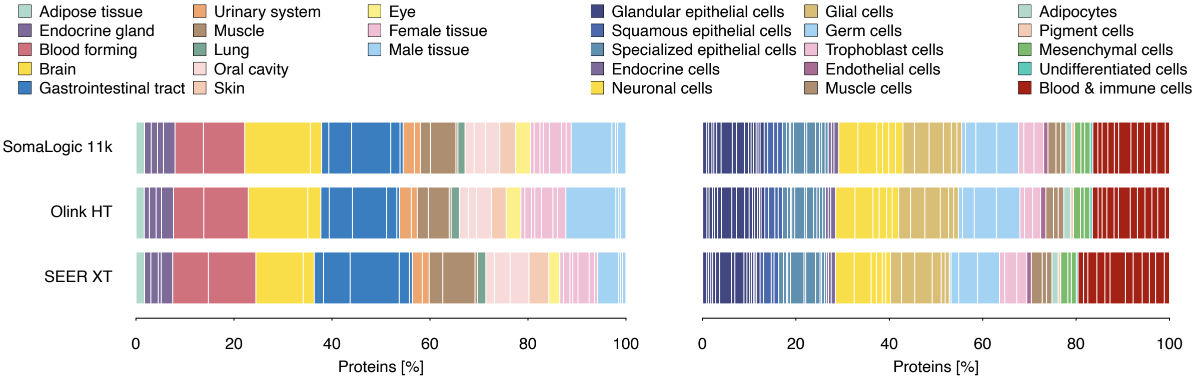


**Supplementary Fig. 2** Coverage of protein targets with evidence for enhanced production in tissues (left) or cell-types (right) inferred from gene expression data based on bulk RNA sequencing in tissues or single cell types. The underlying data was downloaded from the Human Protein Atlas[1].

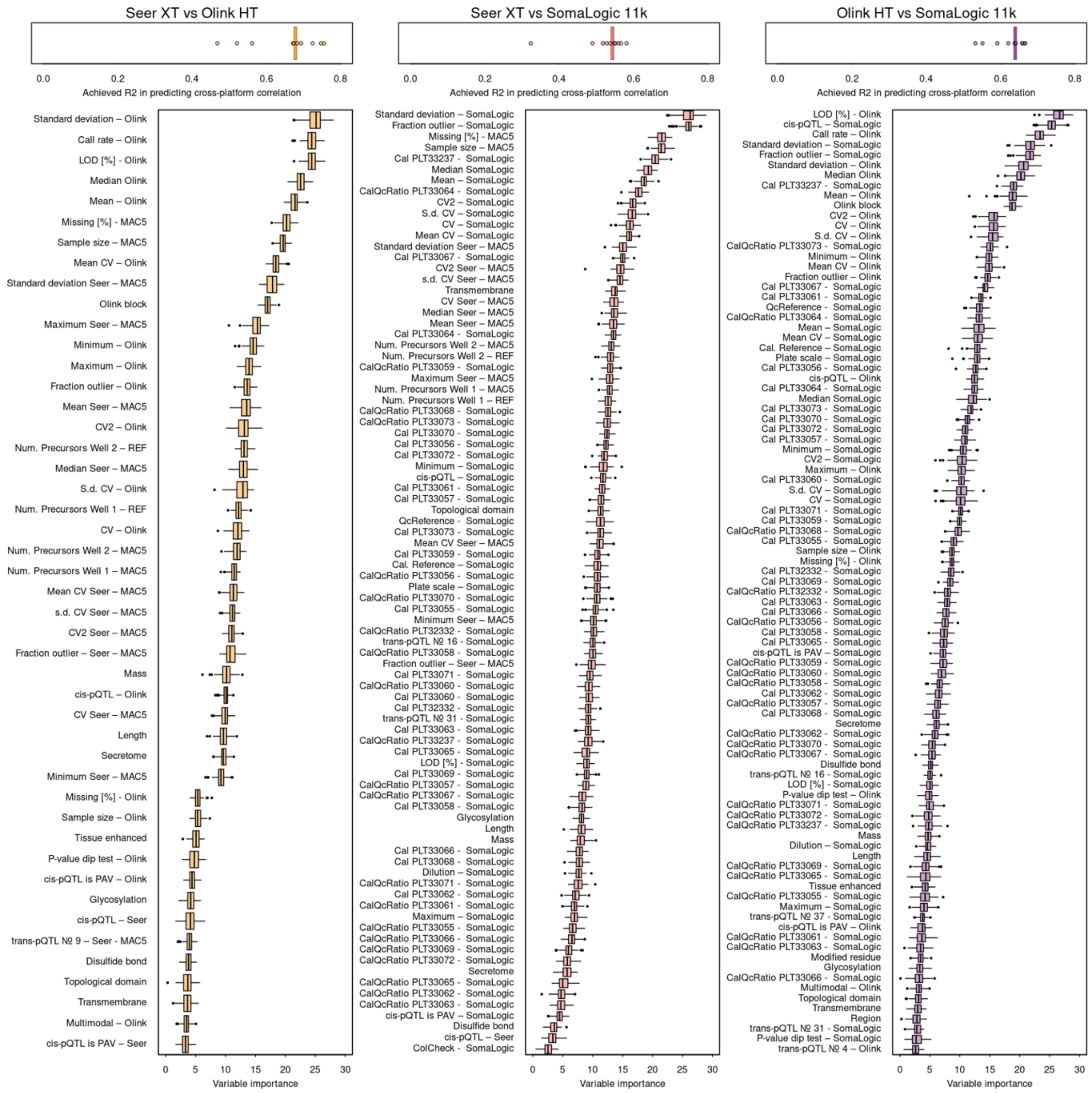


**Supplementary Fig. 3** Model performance (top) and associated feature importance (bottom) for three models (listed on top) to predict the Spearman correlation coefficients for overlapping protein targets. Achieved accuracy is based on a Random Forest classifier trained using 10-fold cross-validation. Feature importance was derived using the Boruta feature selection approach and only variables with confirmed significance ($p<0.05$ following multiple testing correction) are shown. LOD = limit of detection; CV = coefficient of variation; pQTL = protein quantitative trait loci; PAV = protein altering variant

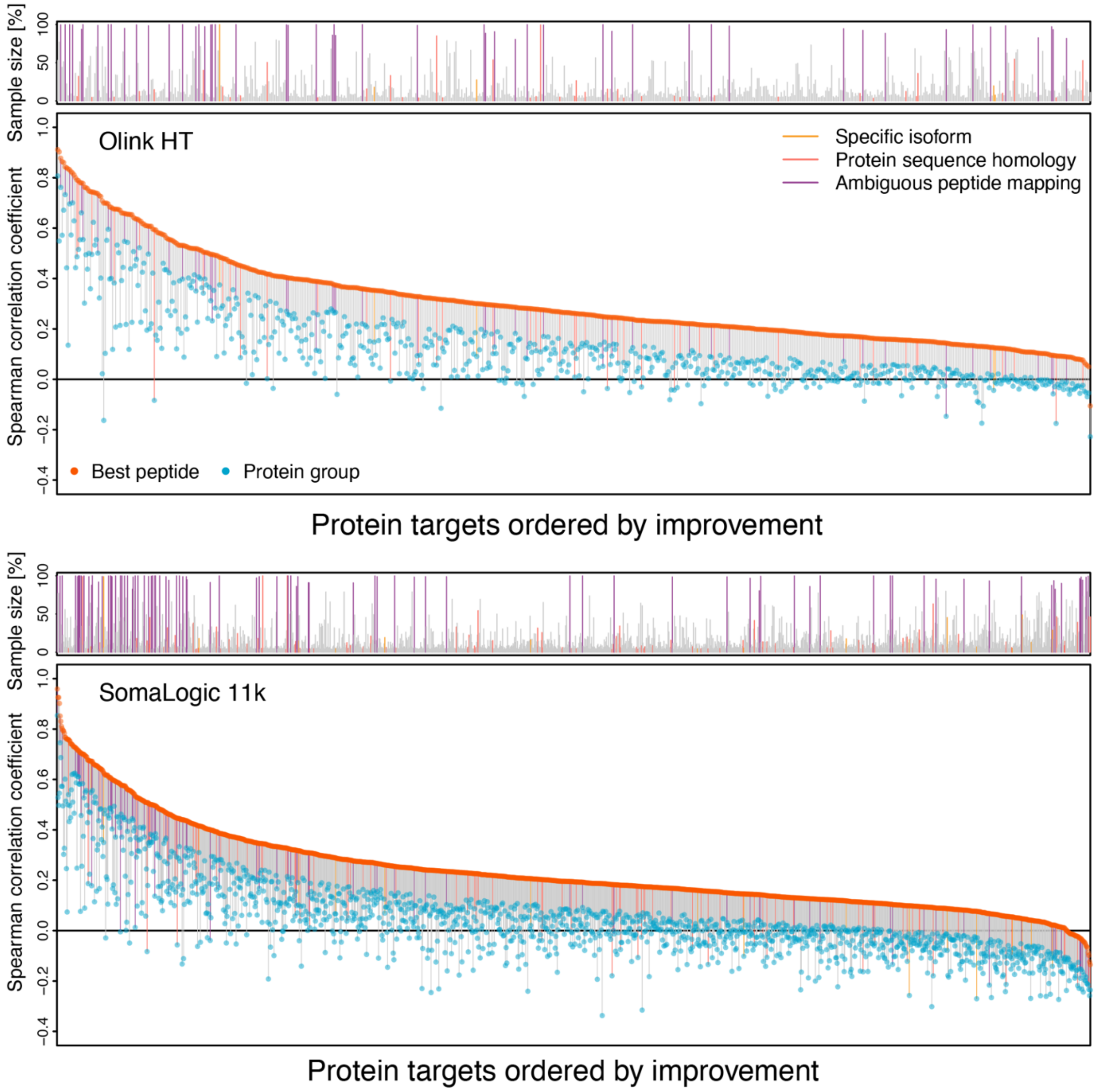


**Supplementary Figure 4** Correlation coefficients that improved >0.1 when comparing measurements for overlapping protein targets based on Olink HT (top) or SomaLogic 11k with peptide level (orange dot) instead of protein group measurements (blue dot). Line charts on top of each plot indicate the sample size for the respective best correlating peptide. Colour code refers to possible explanations for the improvement in correlation coefficients (see Main text).

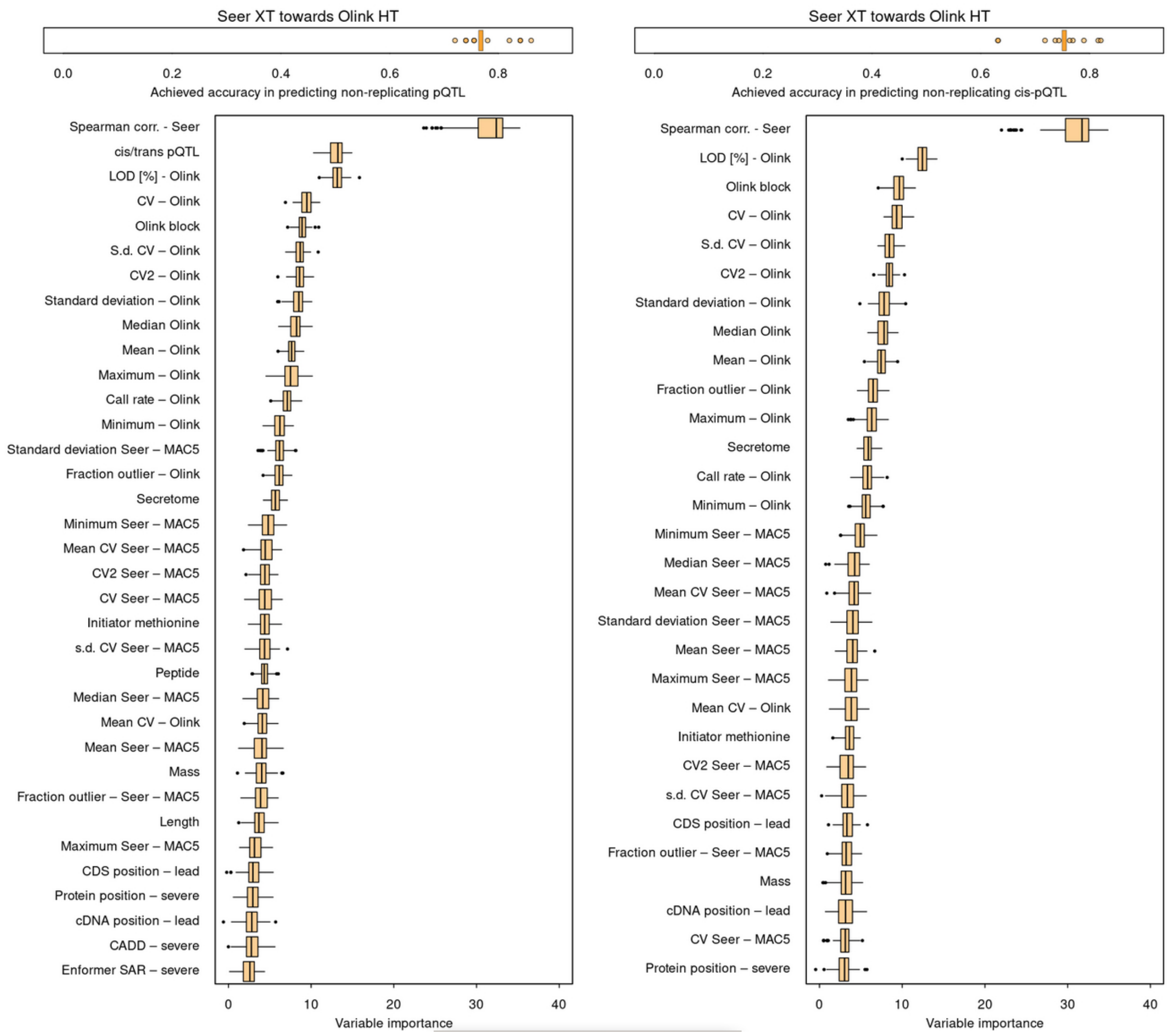


**Supplementary Fig. 5** Model performance (top) and associated feature importance (bottom) for two models (all pQTLs and only cis-pQTLs) to predict replication of pQTLs discovered using Seer Proteograph XT using the Olink HT platform. Achieved accuracy is based on a Random Forest classifier trained using 10-fold cross-validation. Feature importance was derived using the Boruta feature selection approach and only variables with confirmed significance ($p<0.05$ following multiple testing correction) are shown. LOD = limit of detection; CV = coefficient of variation; pQTL = protein quantitative trait loci; PAV = protein altering variant

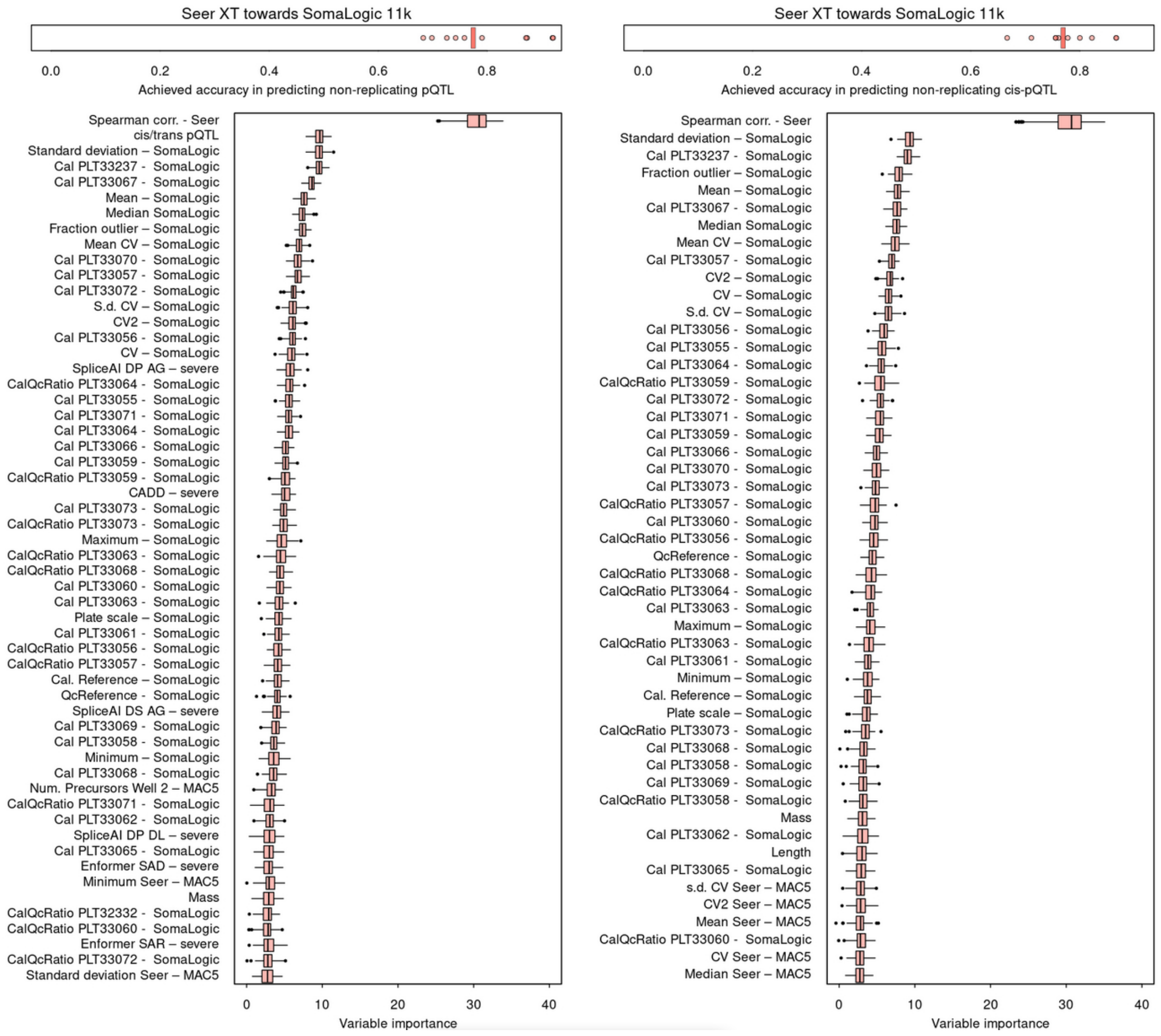


**Supplementary Fig. 6** Model performance (top) and associated feature importance (bottom) for two models (all pQTLs and only cis-pQTLs) to predict replication of pQTLs discovered using Seer Proteograph XT using the SomaLogic 11k platform. Achieved accuracy is based on a Random Forest classifier trained using 10-fold cross-validation. Feature importance was derived using the Boruta feature selection approach and only variables with confirmed significance ($p<0.05$ following multiple testing correction) are shown. LOD = limit of detection; CV = coefficient of variation; pQTL = protein quantitative trait loci; PAV = protein altering variant

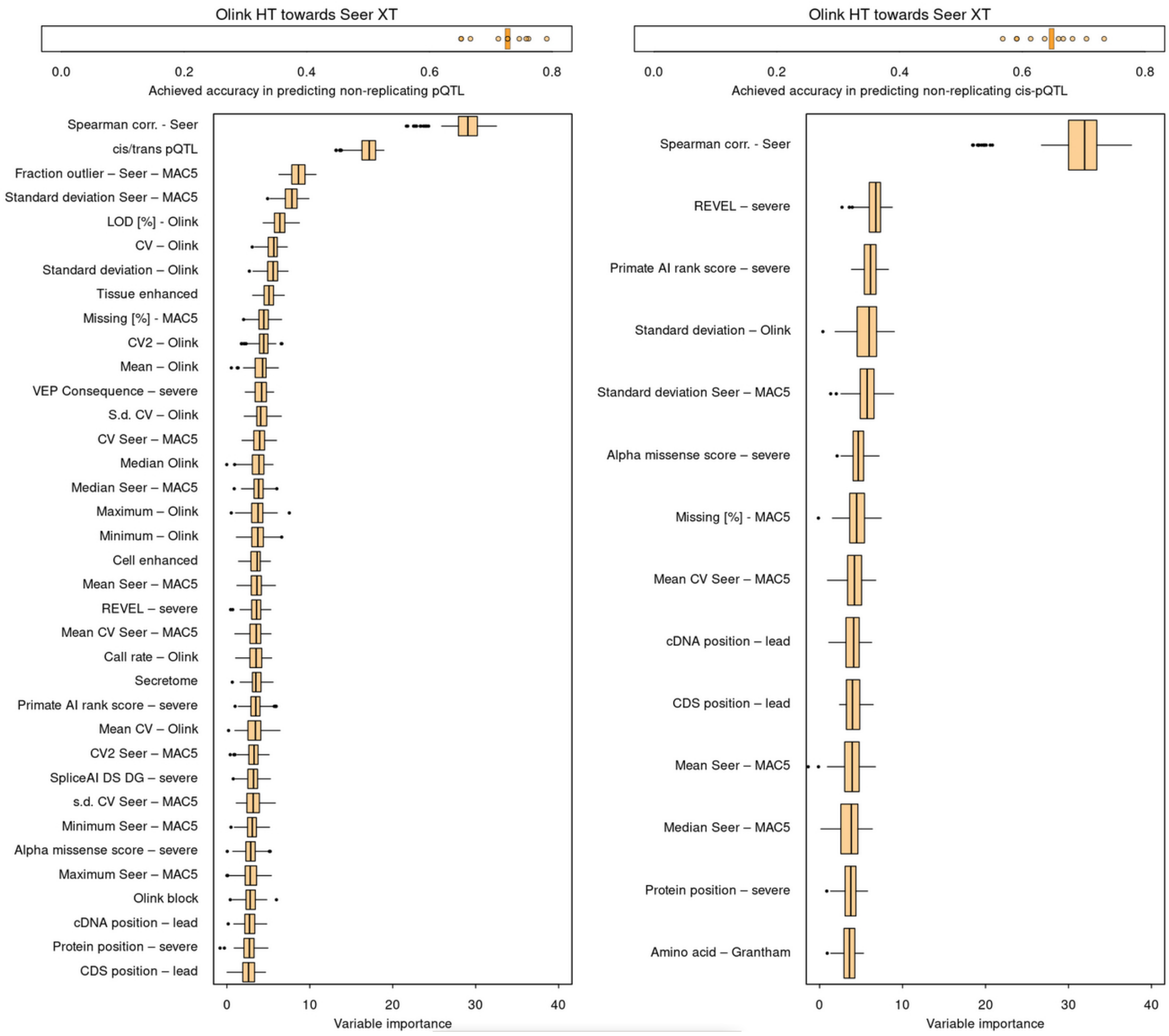


**Supplementary Fig. 7** Model performance (top) and associated feature importance (bottom) for two models (all pQTLs and only cis-pQTLs) to predict replication of pQTLs discovered using the Olink HT by the Seer Proteograph XT platform. Achieved accuracy is based on a Random Forest classifier trained using 10-fold cross-validation. Feature importance was derived using the Boruta feature selection approach and only variables with confirmed significance ($p<0.05$ following multiple testing correction) are shown. LOD = limit of detection; CV = coefficient of variation; pQTL = protein quantitative trait loci; PAV = protein altering variant

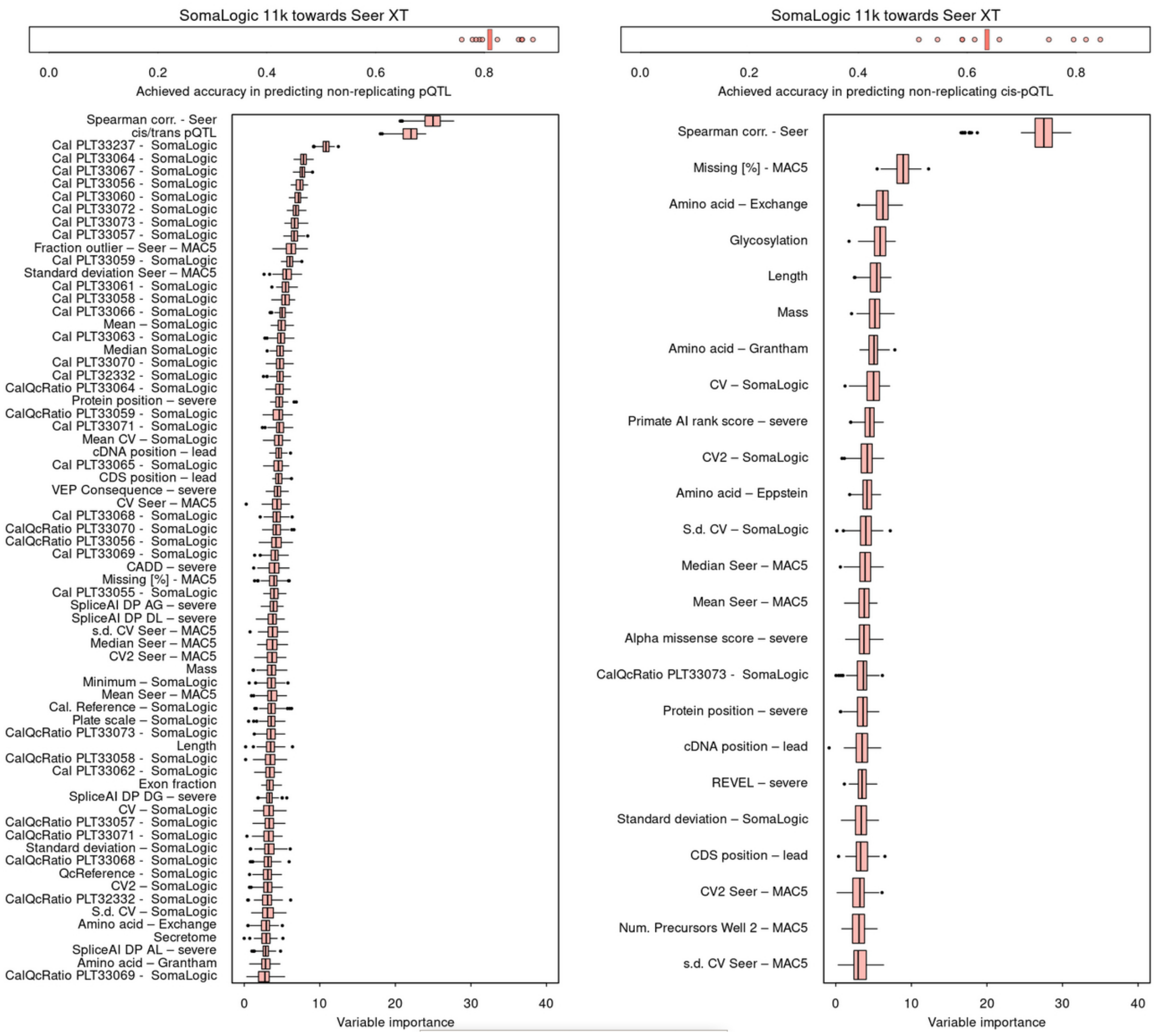


**Supplementary Fig. 8** Model performance (top) and associated feature importance (bottom) for two models (all pQTLs and only cis-pQTLs) to predict replication of pQTLs discovered using SomaLogic 11k by the the Seer Proteograph XT platform. Achieved accuracy is based on a Random Forest classifier trained using 10-fold cross-validation. Feature importance was derived using the Boruta feature selection approach and only variables with confirmed significance (p<0.05 following multiple testing correction) are shown. LOD = limit of detection; CV = coefficient of variation; pQTL = protein quantitative trait loci; PAV = protein altering variant

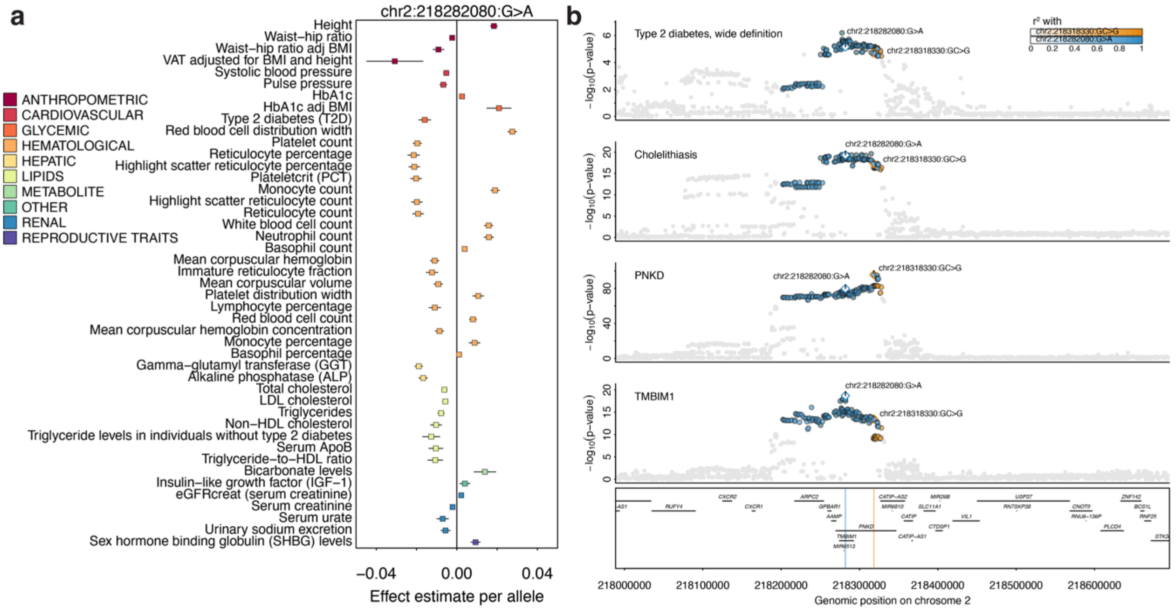


**Supplementary Fig. 9** **a** Forest plot showing significant ($p<10^{-5}$) associations between chr2:218282080:G>A and diverse outcomes obtained from the CMD Knowledge portal (download: 2025/04/15). **b** Stacked regional association plot centred around *TMBIM1* and *PNKD*. The bottom two panels each depict regional association statistics ($-\log_{10}$(p-value)) for plasma levels of transmembrane BAX inhibitor motif-containing 1 (TMBIM1) and PNKD Metallo-Beta-Lactamase Domain Containing (PNKD) from the present study. Association statistics for cholestithiasis and type 2 diabetes were obtained from a meta-analysis of FinnGen and UK Biobank. Variants were coloured based on LD with the respective regional lead variants for each protein. Note that colours overlap for variants in LD with both ($r^2>0.5$; greyish).

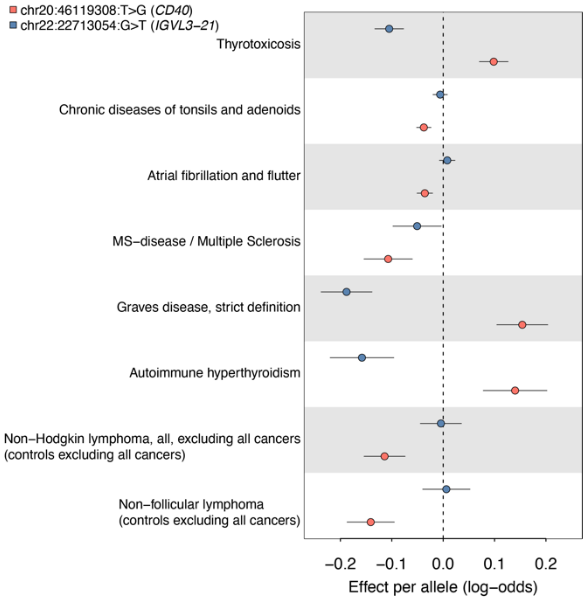


**Supplementary Fig. 10** Forest plot showing effect estimates for two cis-pQTLs (orange: CD40; blue IGLV3-21) across disease with evidence of colocalization for at least one of the two (see Supplementary Tab. 14). Effect estimates were obtained from a meta-analysis of FinnGen and UK Biobank.
